# Using the Behaviour Change Wheel to co-design a sedentary behaviour intervention in individuals with spinal cord injury

**DOI:** 10.64898/2025.12.02.25338957

**Authors:** Daniel L Cooper, Alyson Warland, Emma Norris, Cherry Kilbride, Sue Paddison, Daniel P Bailey

## Abstract

**Purpose:** There is a lack of sedentary behaviour interventions for individuals with spinal cord injury. Interventions designed for non-disabled individuals are unlikely to be generalisable to wheelchair-users with paraplegia, meaning a tailored approach is needed.

**Objective:** This study aimed to co-design a sedentary behaviour intervention using the Behaviour Change Wheel (BCW) for manual wheelchair-users with paraplegia.

**Methods:** An iterative co-design approach was employed across workshops with 10 individuals with paraplegia, 13 healthcare professionals and four community caregivers. Initial workshops focused on barriers and facilitators to reducing and breaking up sedentary behaviour, and possible intervention options. This informed initial intervention concepts, which were mapped to BCW constructs and behaviour change techniques. In follow-up workshops, the acceptability, practicability, effectiveness, affordability, safety/side-effects, and equity of each intervention component was assessed to refine concepts and design an intervention protocol. Data was analysed using the framework method via inductive and deductive coding in context of the BCW.

**Findings:** A multi-component intervention was co-designed and includes (1) a wearable activity tracker to facilitate reminders to be active and give feedback on physical activity, (2) an educational booklet to overcome lack of knowledge around sedentary behaviour, (3) goal setting, (4) one-to-one motivational support from a trained individual, (5) a peer support group and (6) activity tools, including a portable hand cycle and exercise bands.

**Conclusions:** A novel intervention targeting sedentary behaviour has been developed for individuals with paraplegia. The combination of co-design with the BCW likely optimises the intervention’s acceptability and effectiveness, which now requires evaluation.

## Introduction

Individuals with spinal cord injury (SCI) are at significantly greater risk of cardiovascular disease (CVD) than the general population^1^. This increased risk may be a result of physical inactivity, reduced metabolic rate, accumulation of body fat and sarcopenia after injury^2^. Interventions to reduce CVD risk are, therefore, needed in individuals with SCI.

Greater levels of sedentary behaviour, defined as any waking behaviour characterised by an energy expenditure of ≤ 1.5 metabolic equivalents (MET) whilst in a sitting or reclining posture^3^, are associated with an increased risk of CVD, independent of physical activity^4^. Being wheelchair-users through necessity means that most individuals with SCI spend long periods of time being sedentary^5^. Therefore, increased CVD risk in individuals with SCI could also be related to high volumes of sedentary time. A systematic review found a lack of interventions targeting sedentary behaviour in individuals with paraplegia^6^, defined as damage to the spinal cord at the first thoracic vertebrae or below, resulting in trunk and lower limb dysfunction^7^. Most intervention studies included in this review involved structured exercise training^6^, which may not target appropriate behaviour change techniques (BCTs) for reducing sedentary behaviour^8^ and fail to promote non-exercise physical activity across the whole day^9^. This review also identified a lack of behaviour change theory to inform intervention design^6^. Theoretically-driven interventions lead to more positive outcomes^10^, likely due to the recognition of precursors to behaviour and causal factors of change, which can then be selectively targeted with appropriate BCTs^11^. Theoretically-driven interventions are, therefore, needed in individuals with SCI.

Co-design is a process in which key stakeholders play an active role in intervention design and development^12^, resulting in interventions that are more engaging, satisfying and useful to end-users than traditional researcher-developed interventions^13^. Co-design has been used successfully for sedentary behaviour interventions in office workers^14^, older adults^15^ and those with severe mental illness^16^. Embedding co-design within a suitable behaviour change framework, such as the Behaviour Change Wheel (BCW)^17^, has been recognised as a particularly effective method for intervention development^18^. The BCW intervention design process involves an initial behavioural diagnosis using the Capability, Opportunity and Motivation to change Behaviour (COM-B) model and the Theoretical Domains Framework (TDF) to understand the problem and target behaviour^19^. This is followed by identification of intervention functions, policy categories, BCTs and delivery modes through which the intervention will operate. A combination of co-production (a similar technique to co-design) and the BCW was employed to develop a sedentary behaviour intervention for stroke survivors, leading to a feasible and replicable intervention^20^. The use of a combined co-design and BCW approach may, therefore, be an appropriate method for developing a sedentary intervention for individuals with SCI.

Physical activity levels generally increase during inpatient SCI rehabilitation but decline after discharge into the community^21^. This decline is likely due to lack of access to appropriate exercise facilities and the significant challenge of adapting to the home environment after SCI. Although sedentary behaviour improved one year after initial inpatient discharge, levels are still significantly worse compared with non-disabled individuals^5^. Thus, a community-based sedentary behaviour intervention should be designed to support individuals with SCI over the short and longer-term following inpatient rehabilitation.

This study aimed to co-design an intervention, using the BCW framework, to break up and reduce sedentary behaviour in individuals with paraplegia across different stages of the SCI rehabilitation pathway. The objectives were to (a) explore the lived experiences of people with paraplegia regarding barriers and facilitators for breaking up and reducing sedentary behaviour, and (b) identify content and implementation options for the intervention.

## Methods

### Study design and overview

A qualitative workshop approach was utilised to iteratively co-design the intervention, grounded in behaviour change theory using the BCW framework^17^. The co-design process is shown in Figure 1. A total of eight workshops were undertaken separately with participants across three groups: individuals with paraplegia (n = 4 workshops), healthcare professionals (n = 2 workshops), and community caregivers (n = 2 workshops). Workshops were undertaken either online (n = 5 workshops), in-person at Brunel University of London (n = 1 workshop) or at the Royal National Orthopaedic Hospital, London (n = 2 workshops). Activities employed in the workshops included group discussions, writing ideas on post-it notes, visualising ideas using interactive whiteboards, and appraising ideas using Likert rating scales. Figure 2 shows which aspects of the BCW were covered in each workshop. Workshops were facilitated by DLC (MSc; PhD researcher) and supported by AW (PhD), EN (PhD) or DPB (PhD). All researchers had experience in qualitative methods. DLC communicated with participants via email prior to the workshops to organise eligibility screening, consent, and workshop attendance. Introductions were provided by facilitators to explain their role and background at the start of each workshop. The study is reported following the COnsolidated criteria for REporting Qualitative studies^22^.

**Figure 1.**
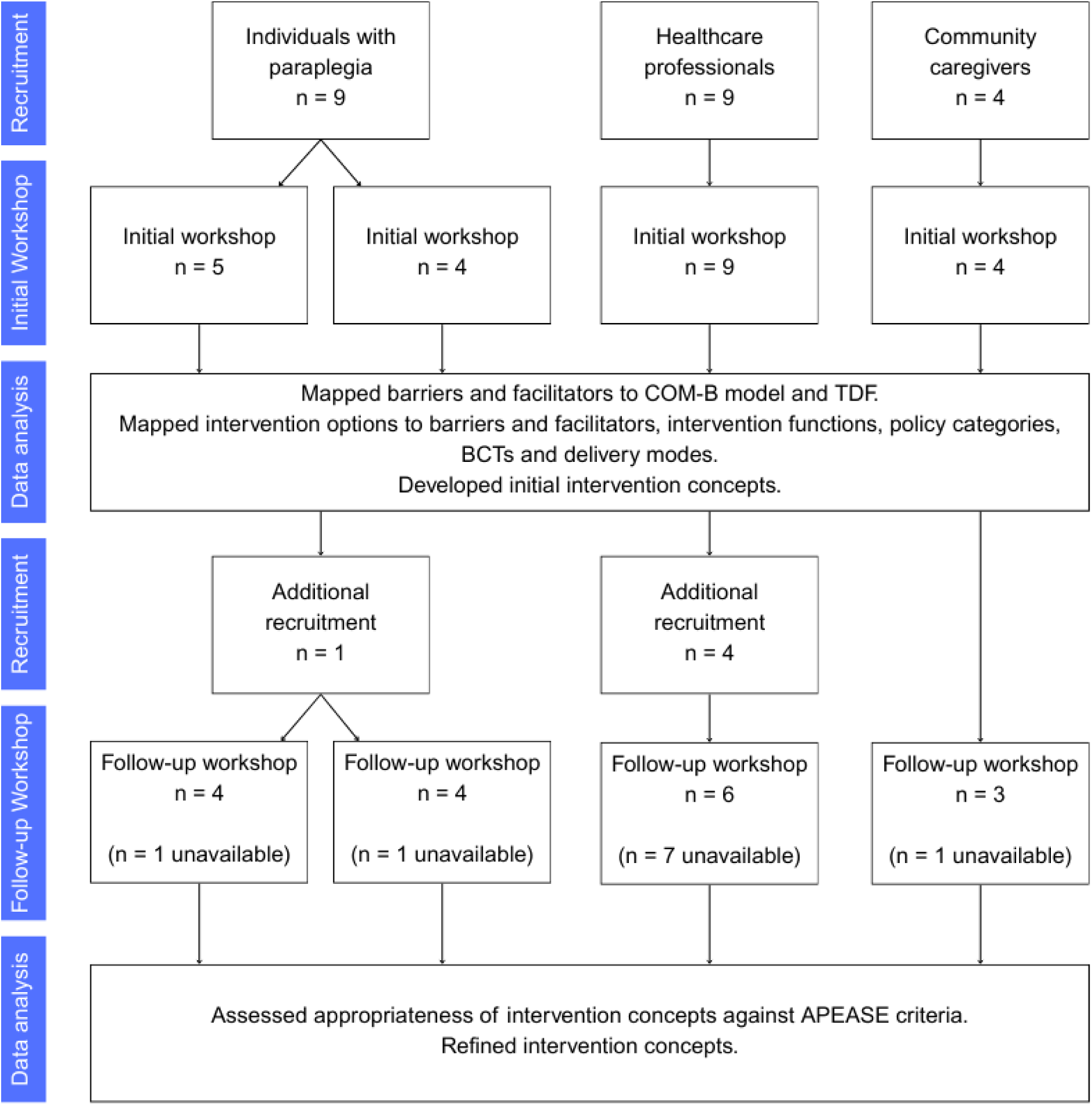
The study co-design process. APEASE, Acceptability, Practicability, Effectiveness, Affordability, Side-effects/safety, Equity; BCT, Behaviour Change Technique; COM-B, Capability, Opportunity and Motivation to change Behaviour; TDF, Theoretical Domains Framework.

**Figure 2.**
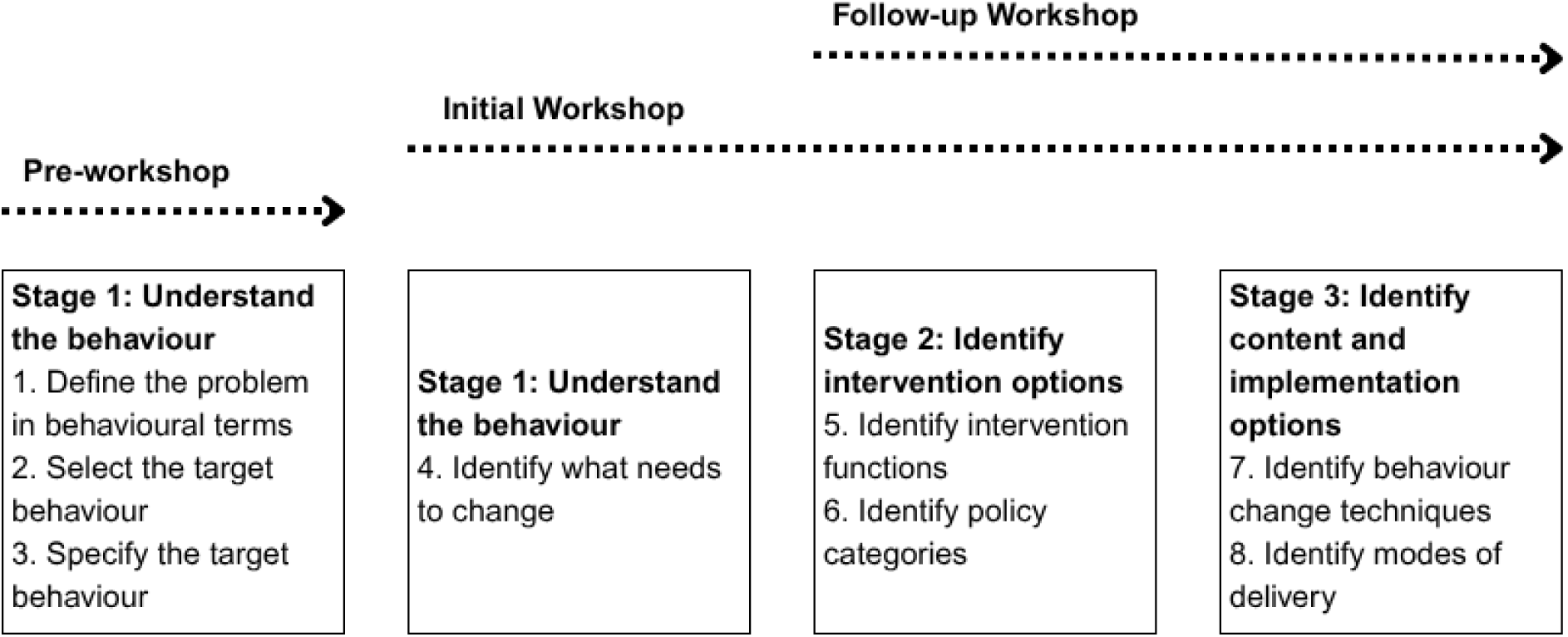
Data collection flowchart detailing the Behaviour Change Wheel stages addressed at each stage of the study

Ethical approval was granted from the College of Health, Medicine and Life Sciences Research Ethics Committee, Brunel University of London (47898-NHS-Apr/2024-50821-2) and the London - Fulham NHS Research Ethics Committee (24/PR/0621). All participants provided informed consent.

### Study sample

Individuals with paraplegia (complete or incomplete SCI) who predominantly used a manual wheelchair for mobility were eligible. Healthcare professionals were eligible if they worked in a hospital or clinic providing care or services for individuals with SCI. Community caregivers provided non-clinical care, services or support to individuals with SCI in the community (friends, family, carers, or employees of relevant organisations/charities), but did not include community healthcare professionals.

### Recruitment

Participants with paraplegia were recruited with the aim of achieving a sample that was representative of individuals from across the SCI care pathway after discharge from initial rehabilitation. This involved recruitment from the community via social media, charity organisations and snowballing, in addition to the London SCI Centre, Royal National Orthopaedic Hospital. Healthcare professionals were also recruited from the London SCI Centre, Royal National Orthopaedic Hospital. Community caregivers were recruited from the community through snowballing and charity organisations.

### Sample size

Research suggests that 6-10 participants per group across 3-12 workshops is sufficient to consider a diverse range of viewpoints and achieve data saturation, whilst still allowing for in-depth discussion^23^. Therefore, a sample size of 6-8 individuals from each participant group was planned for each workshop.

### Intervention development

This study followed the three stages of intervention design outlined in the BCW framework (Figure 2).

#### Stage 1: Understand the behaviour

The problem behaviour, “high sedentary time”, was defined by the research team and discussed with participants at the start of the initial workshops in the context of their experiences around sedentary behaviour. The target behaviour was selected and specified as “reduce and break up sedentary behaviour”.

Identification of what needed to change was achieved by exploring barriers and facilitators to achieving the target behaviour in the initial workshops using the COM-B model as a guide. Barriers and facilitators were mapped to COM-B and the TDF by a researcher after the initial workshops.

#### Stage 2: Identify intervention options

Potential intervention options to address identified barriers and facilitators were proposed by participants during the initial workshops. In line with the BCW framework, each intervention option was then subsequently mapped to intervention functions and policy categories by the research team.

#### Stage 3: Identify content and implementation options

Intervention options were mapped to BCTs from the BCT Taxonomy (BCTT)v1^24^ and modes of delivery by the research team after the initial workshops. This informed the first iteration of intervention concepts. The appropriateness of each concept was appraised using the Acceptability, Practicability, Effectiveness, Affordability, Side-effects/safety and Equity (APEASE) criteria^25^ in follow-up workshops. The content and/or delivery mode of intervention concepts were then refined through appraisal by the research team in collaboration with PPI members.

### Data analysis

Workshops were audio recorded and automatically transcribed using Microsoft Teams (Microsoft Corporation, Redmond, WA, USA). Transcripts were checked for accuracy and participant ID numbers were assigned by a researcher. Data from the initial workshops were analysed by DLC in the context of the BCW using Framework Analysis^26^. Inductive coding was undertaken using NVivo 12 (Lumivero, Burlington, MA, USA) to identify barriers, facilitators and intervention options. Deductive coding followed to map the inductive codes to domains of the BCW and TDF. Credibility in the data was achieved with approximately 20% of initial workshop transcripts being deductively coded by EN. Based on the APEASE appraisal, the research team assessed whether each concept was appropriate for inclusion in the intervention via a rating matrix that determined whether intervention concepts fulfilled each individual APEASE criterion.

Peer debriefing was undertaken with researchers not involved in data analysis throughout the study to challenge assumptions and maintain rigour.

### Patient and public involvement

Two individuals with paraplegia and a healthcare professional contributed to the development of the workshop guides, recruitment methods and participant information sheet. After the workshops, these public contributors reviewed and provided feedback on the appropriateness of the developed intervention materials.

## Findings

### Participant characteristics

Twenty-seven participants took part across the workshops. This included 10 participants with paraplegia (PwP; 4 female); see Table 1 for descriptives, 13 healthcare professionals (HCP; 11 female) and four community caregivers (CCG; two female).

**Table 1.**
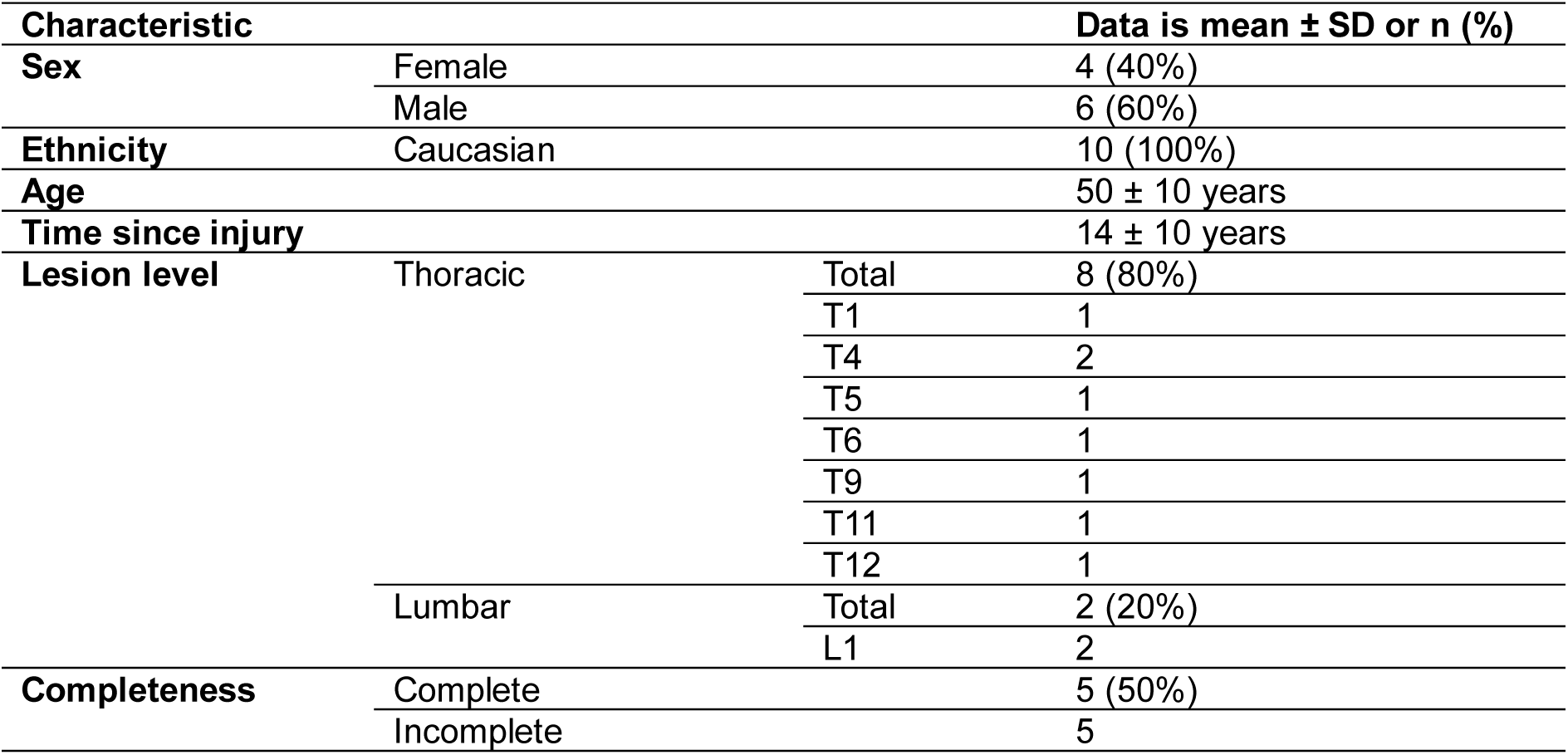
Characteristics of individuals with paraplegia (n = 10).

### Intervention development

Eight workshops were undertaken in total lasting 1 hours 34 minutes on average (range 1-3 hours).

#### BCW stage 1: Understand the behaviour

Barriers and facilitators to reducing and breaking up sedentary behaviour are categorised and reported according to the COM-B model (Table 2).

**Table 2.**
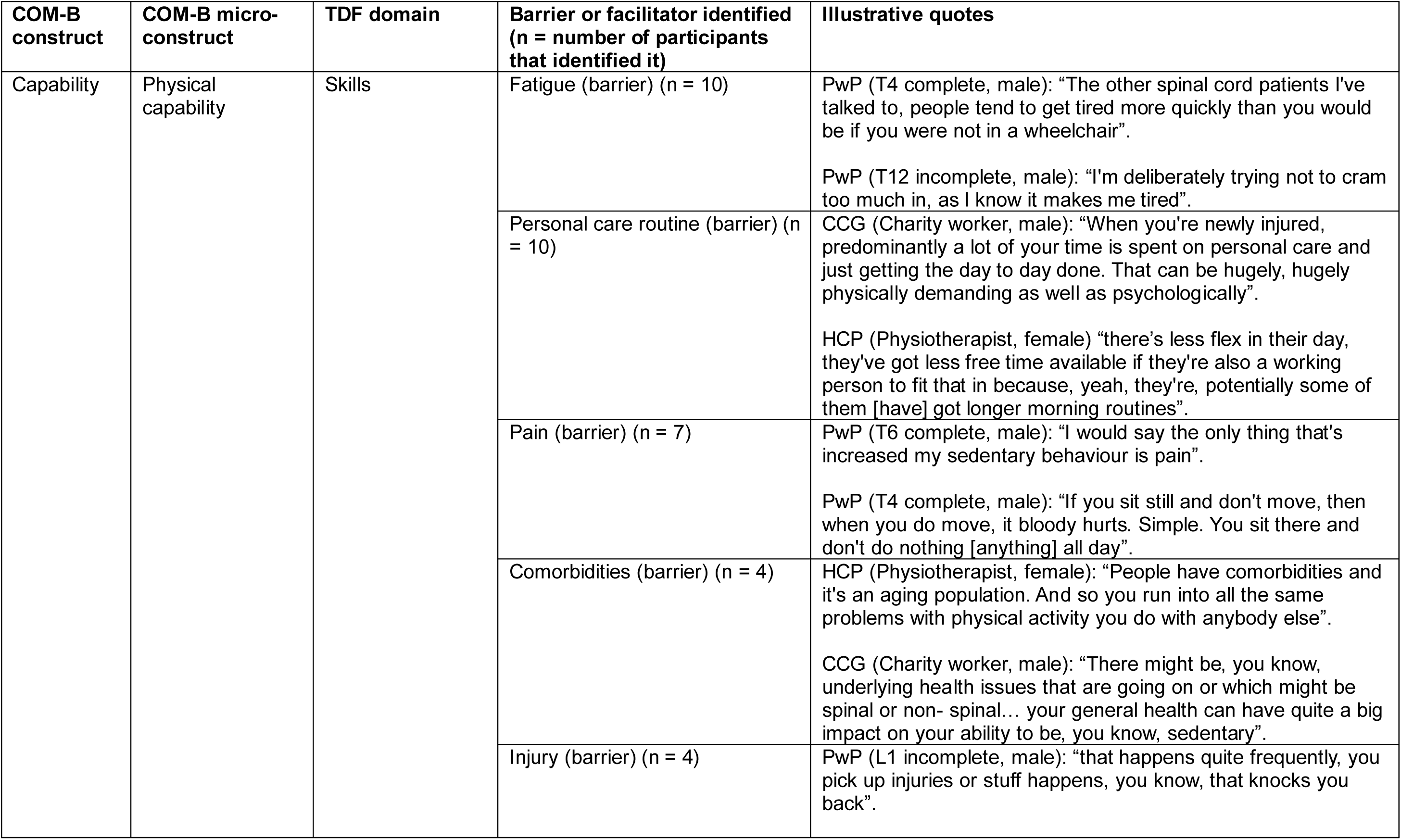
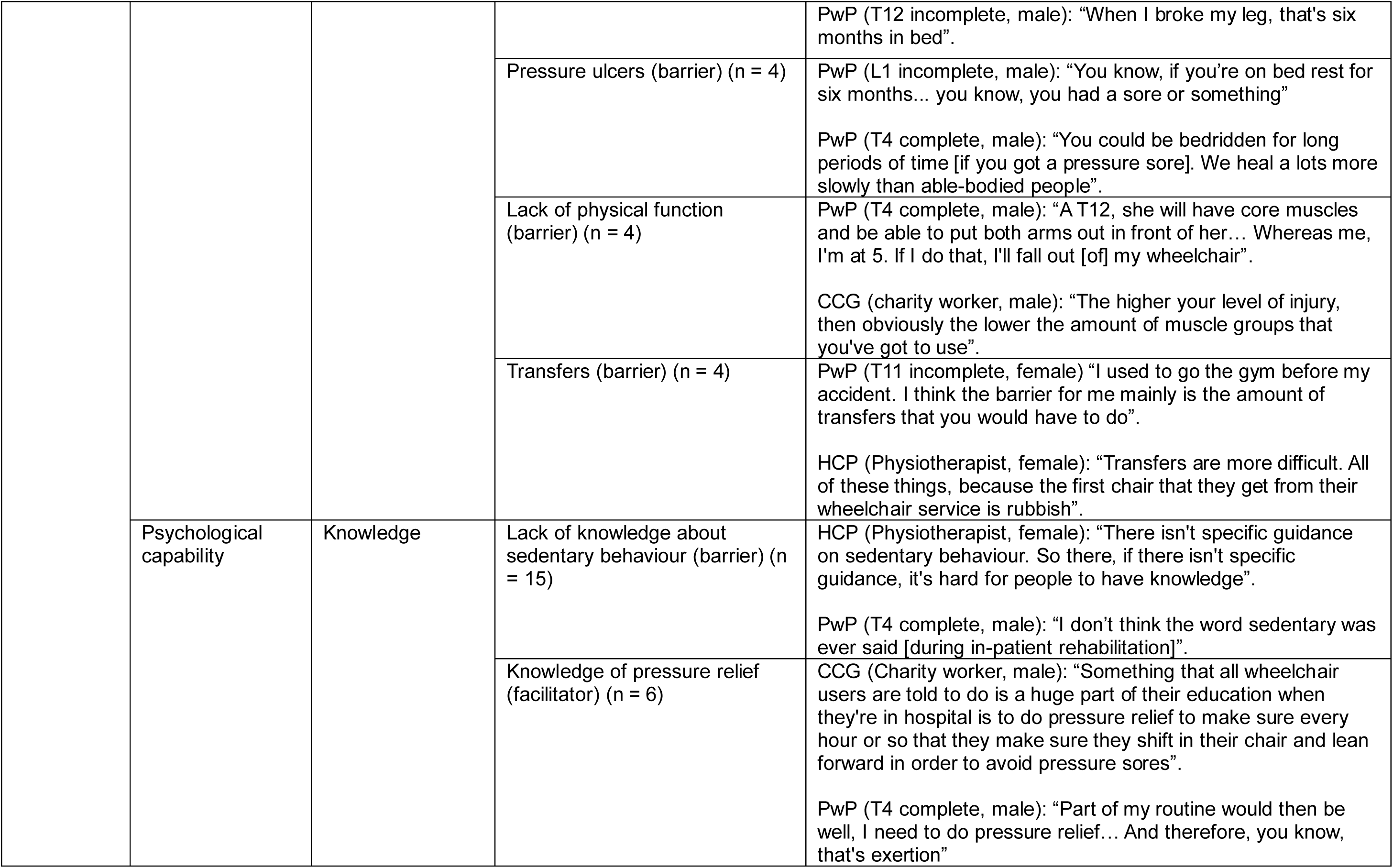
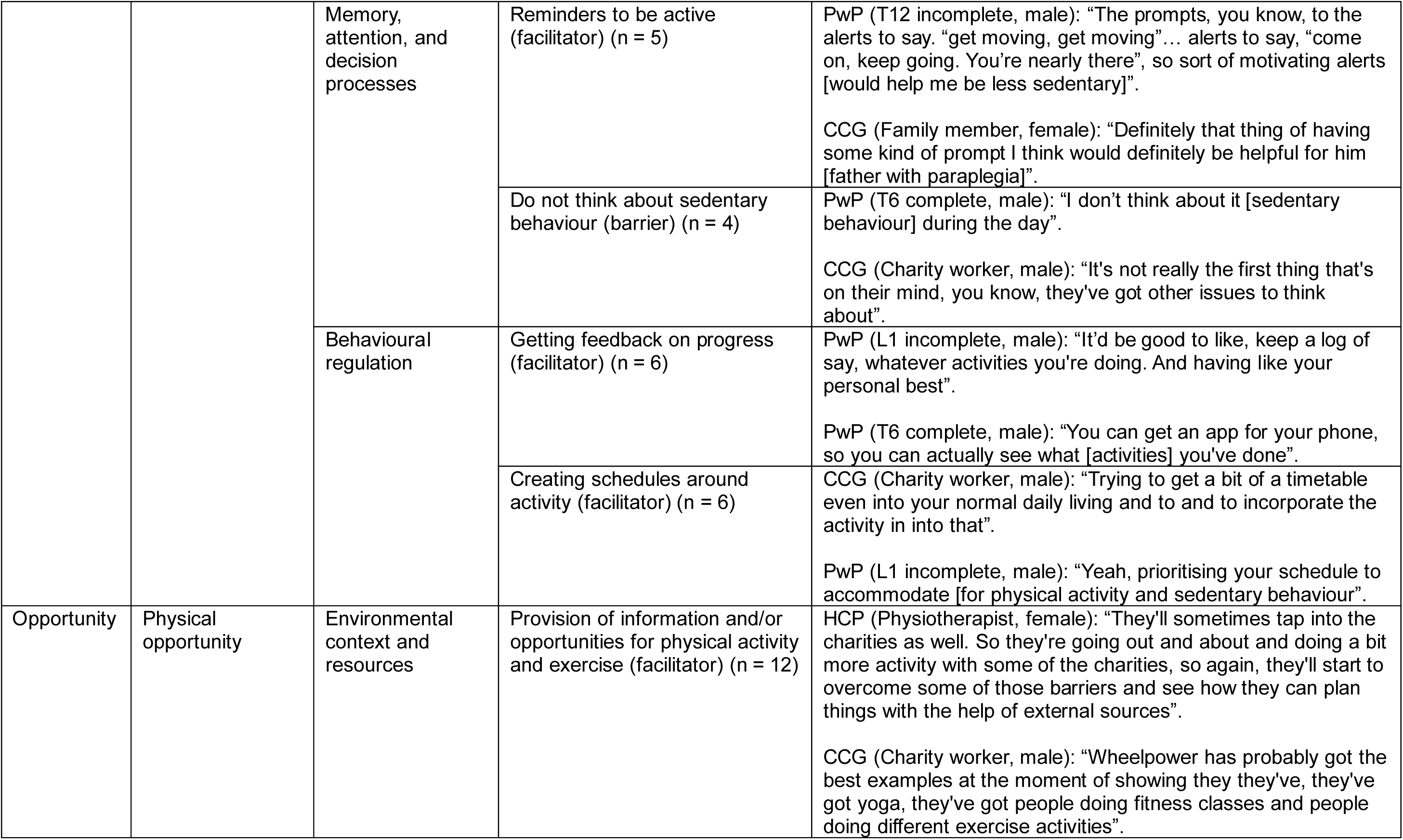
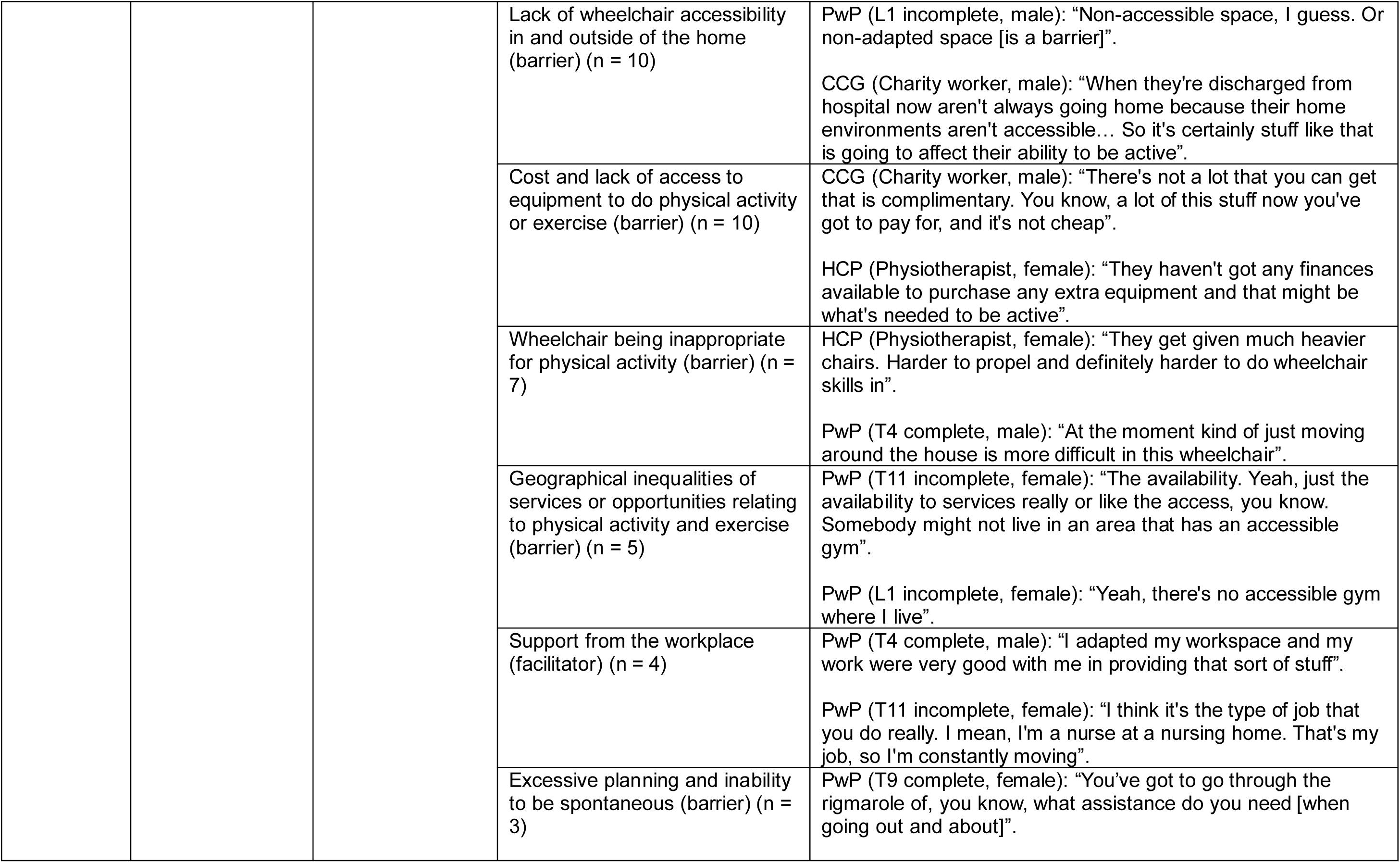
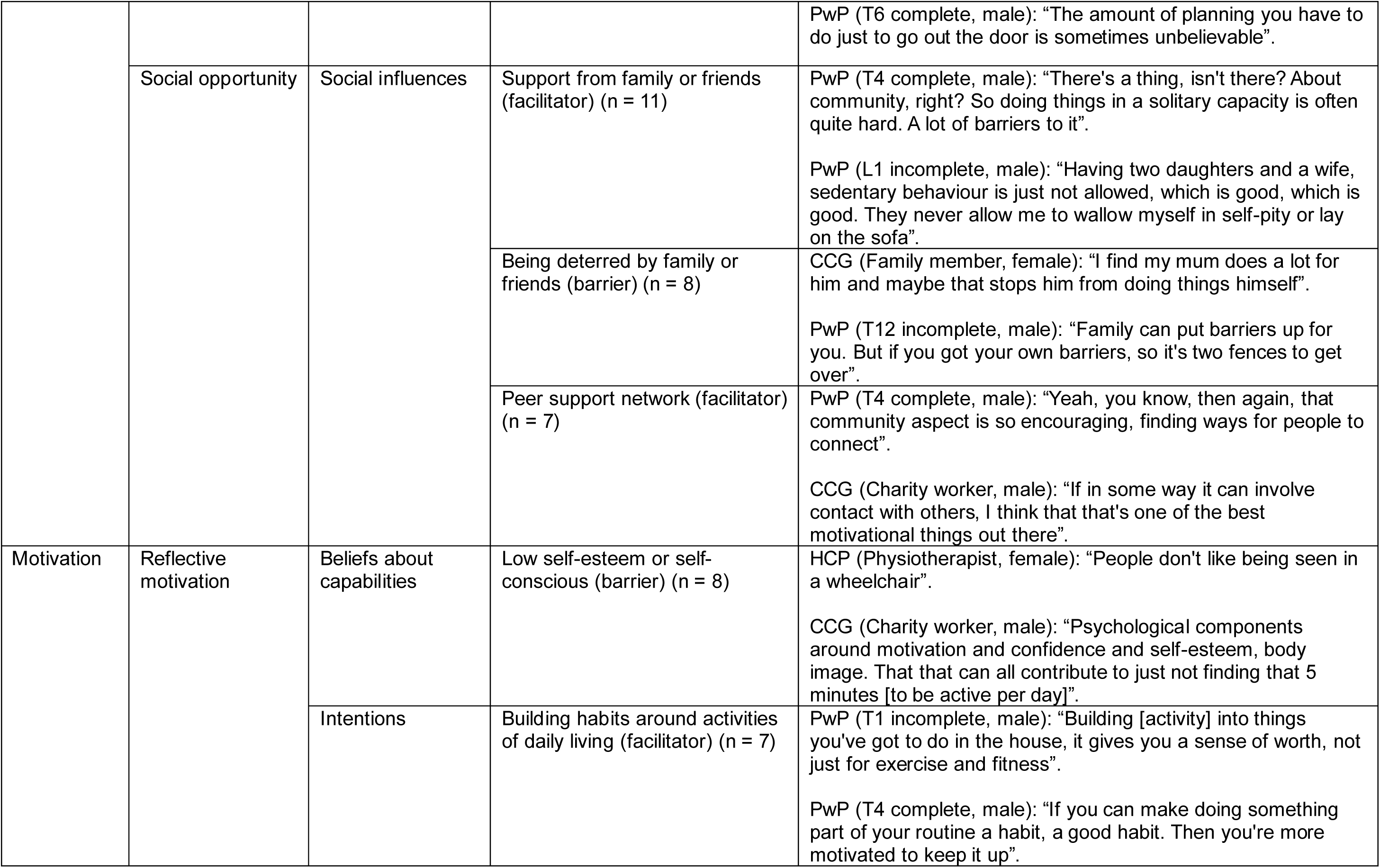
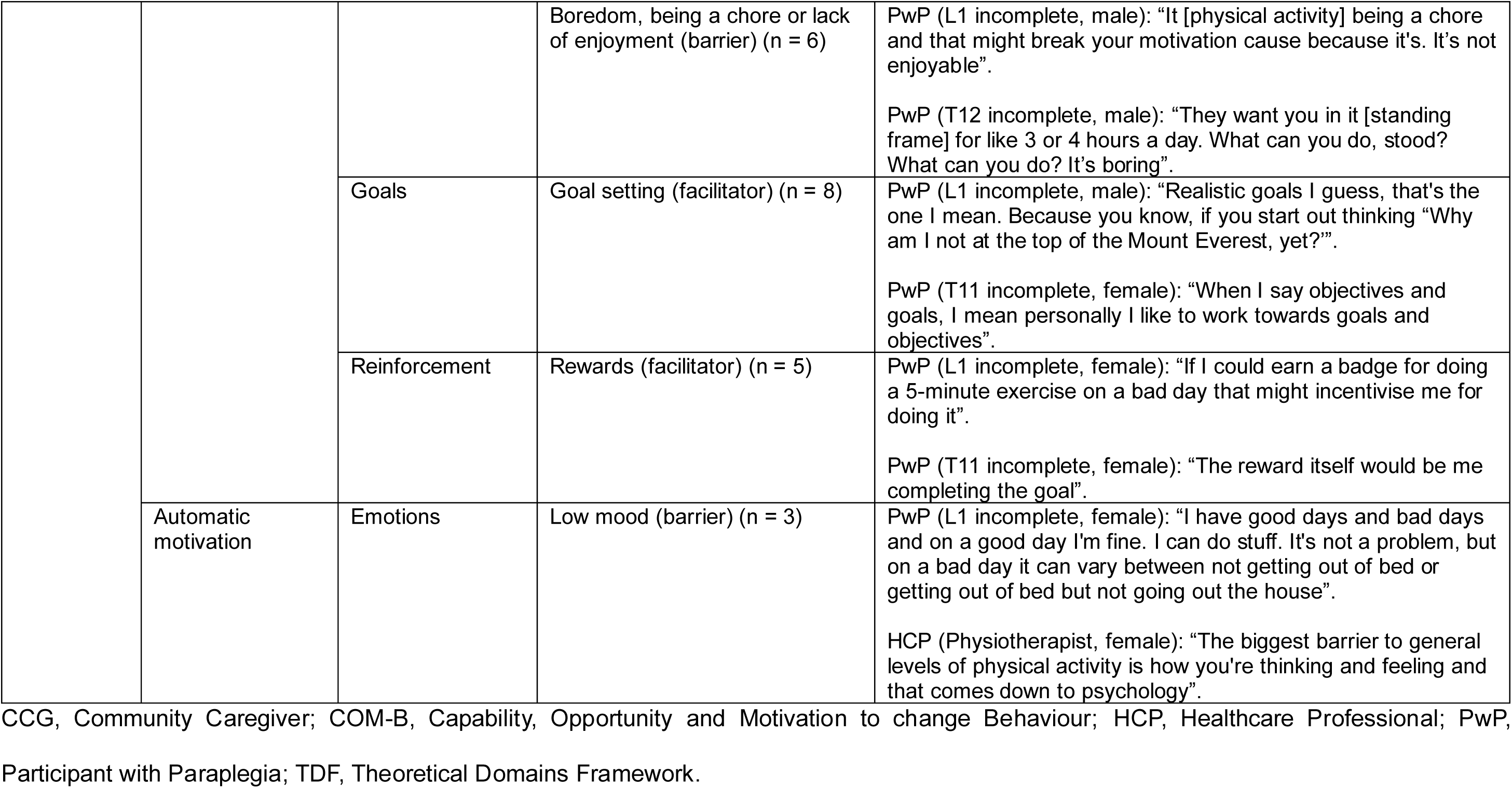
Barriers and facilitators for reducing and breaking up sedentary behaviour.

##### Capability

Common capability-related barriers included lack of knowledge about sedentary behaviour (“*There isn’t specific guidance on sedentary behaviour [in SCI centres]. So, if there isn’t specific guidance, it’s hard for people to have knowledge”* [HCP, Physiotherapist]) and fatigue (*“A lot of your time is spent on personal care and just getting the day-to-days done. That can be hugely, hugely physically demanding”* [CCG, Charity worker]). Personal care routines, pain, comorbidities, injury, pressure ulcers, lack of physical function and necessity to transfer to do physical activity were also common barriers. Facilitators included knowledge of pressure relief, feedback on progress, creating schedules around activity and reminders to be active: *“The text message, with the ‘get up, do something’. Yeah, it works.”* (PwP, T12 incomplete, male).

##### Opportunity

Frequent opportunity-related barriers were lack of wheelchair-accessible space in and outside of the home (“*Non-adapted space is a barrier*” [PwP, L1 incomplete]), cost and lack of access to equipment to do physical activity or exercise (“*There’s not a lot that you can get that is complimentary. You know, a lot of this stuff you’ve got to pay for and it’s not cheap*” [CCG, Charity worker]), being deterred by family and friends, wheelchairs being inappropriate for physical activity, and geographical inequalities of services or opportunities relating to physical activity and exercise. Facilitators included provision of information and/or opportunities for physical activity and exercise, support from family or friends, and a peer support network: *“There’s a thing, isn’t there? About community, right? So doing things in a solitary capacity is often quite hard. A lot of barriers to it”* (PwP, T4 complete).

##### Motivation

Motivation-related barriers included low self-esteem or self-consciousness (*“People don’t like being seen in a wheelchair”* [HCP, Physiotherapist]), boredom, being a chore or lack of enjoyment, and low mood. Facilitators included goal setting (*“I like to work towards goals and objectives. The reward itself would be me completing the goal”* [PwP, T11 incomplete]), building habits around activities of daily living (*“If you can make doing something part of your routine a habit, a good habit, then you’re more motivated to keep it up”* [PwP, T4 complete]) and rewards.

#### BCW stage 2: Identify intervention options

Ten different intervention options were identified (Table 3), including (1) receiving feedback on sedentary behaviour and physical activity, to help facilitate feedback on progress:*“[It would] be good to like, keep a log of say yeah, whatever activities you’re doing. And having, like, your personal best”* (PwP, L1 incomplete). Reminders to break up sedentary behaviour (2) was commonly proposed to address not thinking about sedentary behaviour (barrier): “*Having some kind of prompt [to get moving] I think would definitely be helpful for him*” (CCG, Family member). Education (3) was suggested to address lack of knowledge about sedentary behaviour (barrier): *“People really need to be made aware of what that [reducing sedentary behaviour] looks like, what that could be, and also and also what the benefits are physiologically and psychologically”* (CCG, Charity worker). To address cost and lack of access to equipment to do physical activity or exercise (barrier), having physical activity equipment (4) was identified as an option: *“They haven’t got any finances available to purchase any extra [exercise] equipment and that might be what’s needed to be active”* (HCP, Physiotherapist).

**Table 3.**
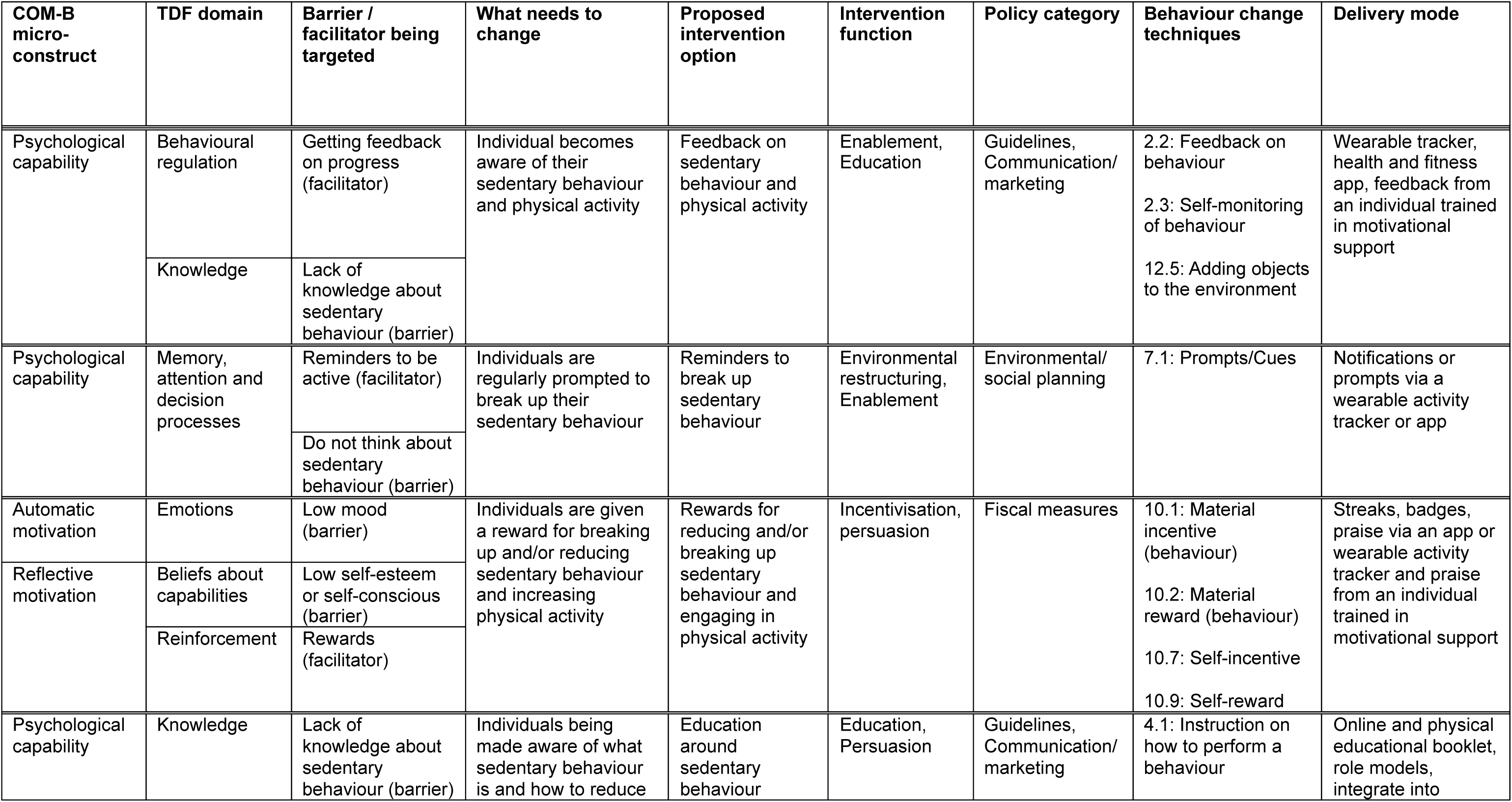
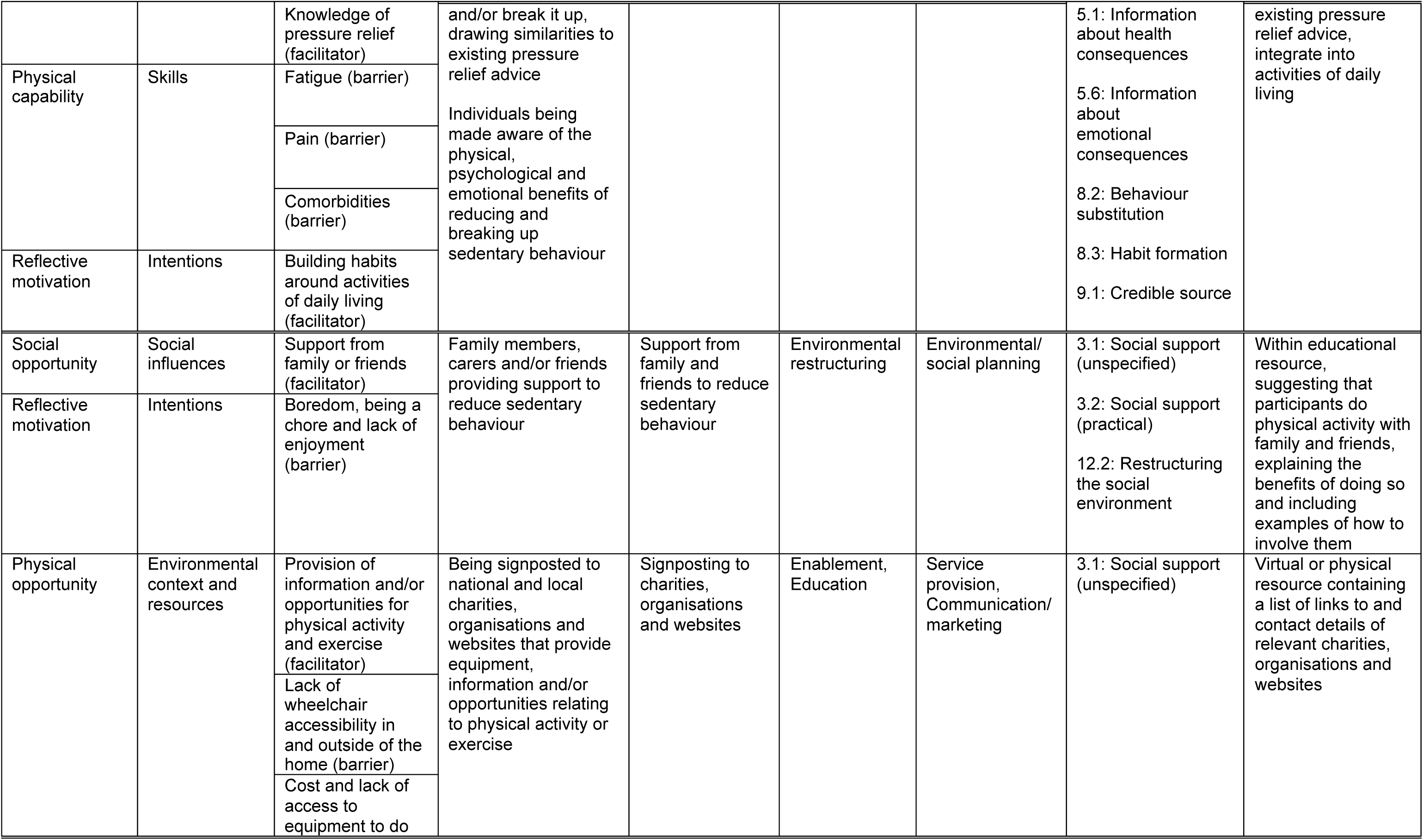
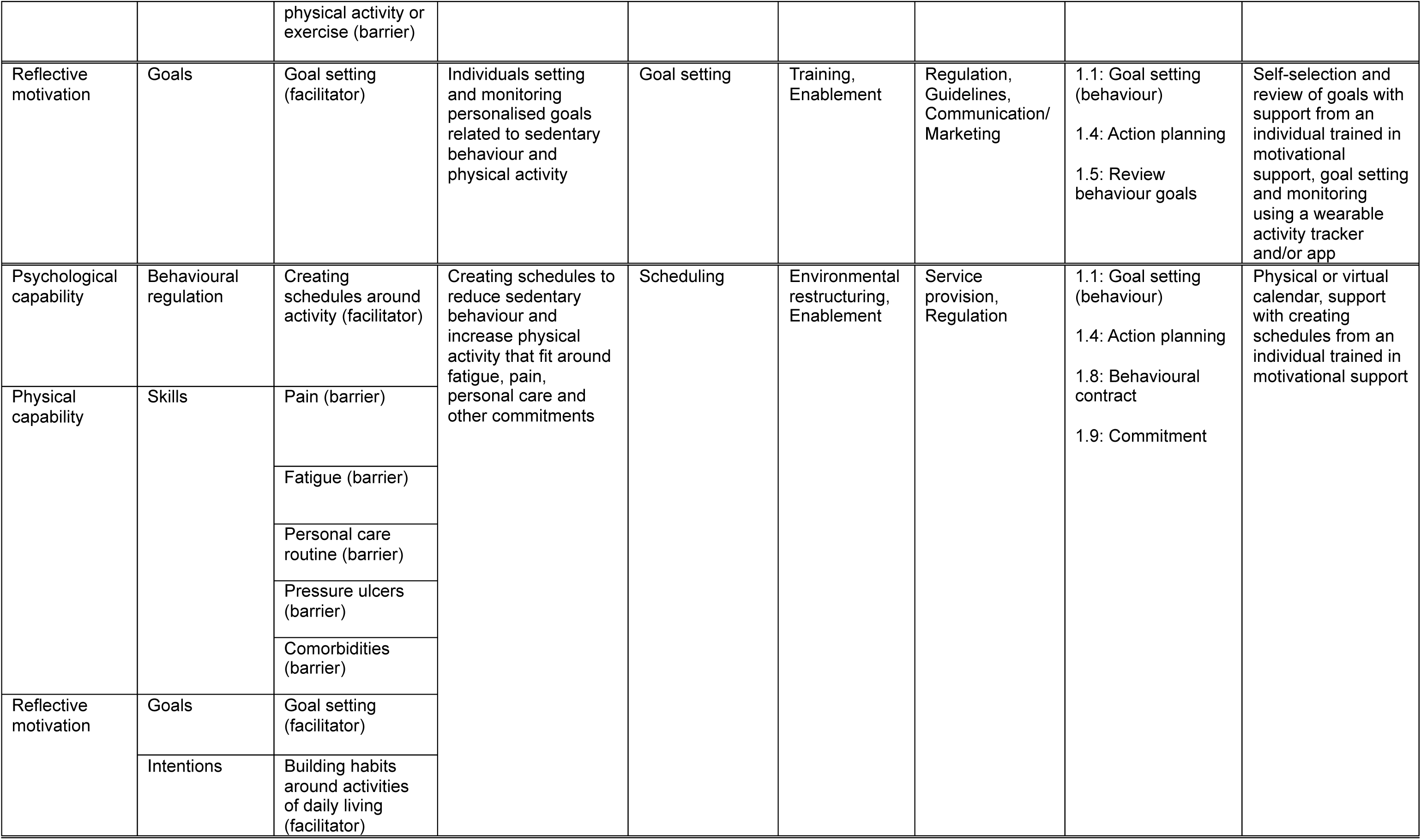
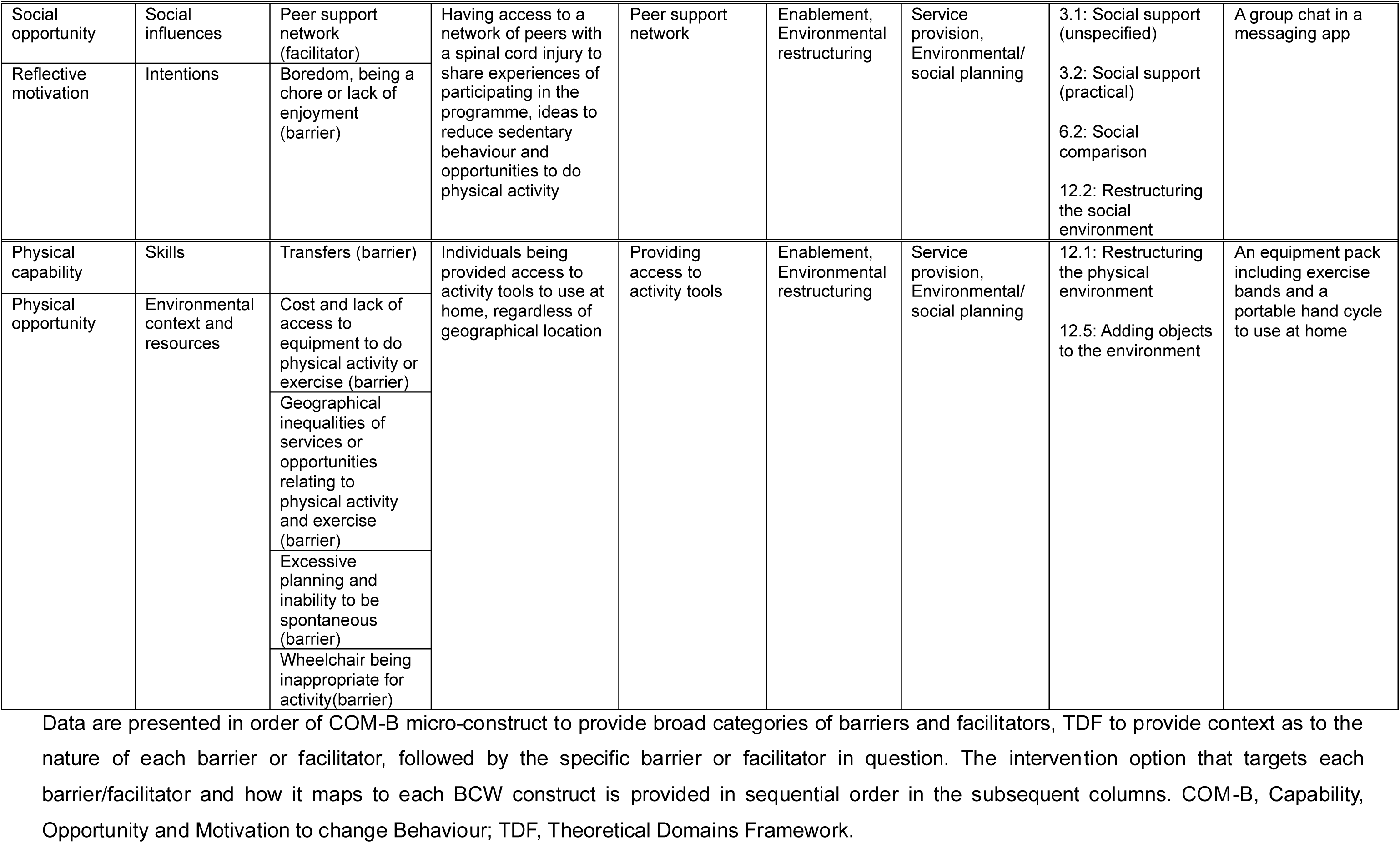
Intervention options mapped to the COM-B, TDF and BCW constructs.

A peer support network was suggested as an option to facilitate peer support (5): *“If you’re feeling a little bit demotivated, you know, maybe there’s like a chat or a forum or something”* (PwP, T6 complete). Getting support from family and friends (6) was also suggested to facilitate peer support: *“I think it’s good to involve family and friends. If they’re up for doing it like people have already said, I think it’s a good motivation”* (CCG, Charity Worker). Goal setting (7) came through as an intervention option: *“Could there be like a mentoring or coaching? It could be part of the creating targets. Maybe you can check back?”* (PwP, L1 incomplete). Creating schedules (8) was an option to support planning around activity (facilitator): *“Trying to get a bit of a timetable even into your normal daily living and to and to incorporate the activity in into that”* (CCG, Charity Worker). Signposting to relevant organisations (9) would facilitate the provision of information and/or opportunities for physical activity and exercise: “*If you have a website, you can link it to the local sports groups*” (PwP, T1 incomplete). Gaining rewards for reducing sedentary behaviour (10) could address low confidence or self-esteem (barrier): *“If I could earn a badge for doing a 5-minute exercise on a bad day that might incentivise me for doing it*” (PwP, L1 incomplete).

##### Identification of intervention functions and policy categories

Six of nine BCW intervention functions were identified as relevant to the intervention options (Table 3). These were Education (n = 3 intervention options were mapped to this function), Enablement (n = 7), Environmental Restructuring (n = 5), Incentivisation (n = 1), Persuasion (n = 2) and Training (n = 1). Six of seven policy categories in the BCW guide were identified as relevant (Table 3), namely Communication/Marketing (n = 4), Environmental/Social Planning (n = 4), Fiscal Measures (n = 1), Guidelines (n = 3), Regulation (n = 2) and Service Provision (n = 4).

#### BCW stage 3: Identify content and implementation options

Twenty-four BCTs were mapped to the intervention options (Table 3) from the following BCTTv1^24^ categories: Goals and Planning, Feedback and Monitoring, Social support, Shaping Knowledge, Natural Consequences, Comparison of Behaviour, Associations, Repetition and Substitution, Comparison of Outcomes, Reward and Threat, and Antecedents.

A wearable activity tracker with messaging relevant to wheelchair-users was suggested for providing feedback on physical activity, reminders to break up sedentary behaviour and virtual rewards for breaking up sedentary behaviour and engaging in physical activity. It was proposed that education around sedentary behaviour, signposting to relevant organisations, and information on support from family and friends could be delivered via an educational booklet. Proposed delivery modes for goal setting and creating schedules were a goal-setting worksheet and one-to-one support from a person trained in motivational interviewing. A social messaging app group chat was proposed to facilitate peer support. A pack of activity tools comprising exercise bands and a portable hand cycle was proposed (access to physical activity equipment). Intervention options with overlapping modes of delivery were merged into six intervention components by the research team after the initial workshops and subsequently discussed with participants in follow-up workshops to assess appropriateness and inform refinements.

##### APEASE assessment

A wearable activity tracker was deemed likely to be effective, with no concerns raised around acceptability, practicability, or safety (Table 4). Concerns were raised around affordability. Equitability was questioned in relation to individuals less proficient with this type of technology but it was felt this could be overcome if participants were provided with guidance.

**Table 4.** Assessment of intervention components using the APEASE criteria.

| Intervention component | Acceptability | Practicability | Effectiveness | Affordability | Safety/side effects | Equity | Comments |
| --- | --- | --- | --- | --- | --- | --- | --- |
| **Wearable activity tracker** | ✓ | ✓ | ✓ | X | ✓ | ✓ | It may not be affordable if participants were required to purchase wearable trackers with wheelchair mode. Concerns around the technological proficiency required are overcome by researchers providing guidance. |
| **Educational booklet** | ✓ | ✓ | ✓ | ✓ | ✓ | ✓ | Concerns around accessibility of the format are overcome by offering both printed and electronic versions. Concerns around acceptability of terminology overcome by using inclusive and positive wording in the materials. |
| **Goal setting** | ✓ | ✓ | ✓ | ✓ | ✓ | ✓ | Concerns around lack of support with creating and working towards goals are overcome by the inclusion of motivational support. |
| **Motivational support** | ✓ | ✓ | ✓ | ✓ | ✓ | ✓ | Concerns around the practicability of access to a medically qualified individual are overcome through delivery by an individual with suitable motivational support training. |
| **Peer support group** | X | ✓ | ✓ | ✓ | X | ✓ | Concerns around acceptability of social comparison are overcome by not sharing activity data between participants. Concerns around privacy are overcome by this component being optional and setting clear ground rules. |
| **Activity tools** | ✓ | ✓ | ✓ | ✓ | ✓ | ✓ | Safety concerns around injury are overcome by providing guidance for using the tools safely. |

An educational booklet was deemed likely to be effective. No concerns were raised regarding practicality, affordability or safety. Printed and electronic formats were recommended for ease of access. The term ‘sedentary’ was not deemed acceptable due to the stigma attached to wheelchair use: “*Sedentary means you sit on your bum all day. We don’t have a choice. So, I would say ‘inactivity’, ‘improve activity’ or something more positive, rather than leaning on the negative*” (PwP, T1 incomplete). Therefore, the term ‘inactive’ was suggested. It was suggested that positive messaging should be used where possible, (e.g. “break up inactivity”, “do more activity breaks”) rather than limiting sedentary behaviour (e.g. “reduce inactivity”).

Participant feedback on goal setting was generally positive with acceptability, effectiveness, affordability and safety not raising any concerns. Regarding practicability and equity, it was proposed that some participants may require more support than others. This led to the suggestion that motivational support sessions with a trained person could support goal setting through accountability and social support.

Peer support was generally deemed acceptable, practicable, effective, safe and equitable. Most participants noted a widely used app (i.e. WhatsApp) would be unlikely to exclude many participants. However, concerns about the privacy of a group chat were raised in addition to concerns about the effectiveness of such a group as some participants did not feel that they needed peer support. Social comparison of progress with the intervention was deemed less acceptable. Healthcare professionals raised concerns around the risk of undesirable behaviour in the group. To address these concerns, it was agreed that this component of the intervention should be optional, have ground rules for using the chosen app and social comparison should not be advocated.

The activity tools were generally considered to be effective. Some participants suggested tools could be difficult to use due to a lack of knowledge for their use or risk of injury. Therefore, a guidance document was added to this component alongside links to demonstrative videos in the educational booklet.

## Discussion

This co-design study has led to the development of a novel intervention targeting sedentary behaviour in individuals with paraplegia. A rigorous co-design approach embedded within the BCW was adopted to maximise the intervention’s effectiveness, feasibility and acceptability. The rigorous combined approach and the novelty of the intervention advances knowledge related to developing behavioural interventions and addressing sedentary behaviour in individuals with paraplegia.

The co-design approach employed was acceptable, with high levels of engagement and retention. The opportunity to speak with peers about common problems and potential options to overcome them within the workshops was noted as a positive experience. Co-design may have generated greater participant engagement than traditional researcher-led methods due to a sense of ownership arising from equal partnership^27,28^. Combining co-design with a workshop-based approach may have increased peer discussion, trust, and openness between participants^29^, integral to sharing richer and more honest insights^29^. The co-design workshop approach, therefore, led to a rich dataset and targeted intervention components appropriate for individuals with paraplegia.

The BCW was adopted in this study due to its comprehensive, theory-driven approach to intervention development^17^. While the co-design workshops allowed exploration of participant barriers and facilitators, the BCW facilitated a coherent behavioural diagnosis^25^. As the co-design approach facilitated discussion of preferred intervention options, the BCW allowed the systematic mapping of these^25^, meaning decision-making can be traced transparently. The APEASE assessment ensured intervention concepts are feasible, acceptable and safe before resources are spent on testing them. The use of behaviour change theory can lead to greater behaviour change in individuals with physical disabilities^10^. Therefore, it is likely that the rigorous combined co-design and BCW will result in a more acceptable and effective intervention.

The co-design process led to the need for a multi-component intervention to address a range of unique barriers and facilitators for individuals with paraplegia. This is in line with research demonstrating the superiority of multi-component interventions for reducing sedentary behaviour over single-component interventions in clinical populations^30^. Additionally, the inclusion of a wrist-worn, wearable activity tracker is consistent with interventions that have reduced sedentary behaviour in non-disabled individuals^31,32^ and clinical populations^30^. Several BCTs identified in the present study, such as self-monitoring, social support, feedback, and prompts and cues, have been effective in previous sedentary behaviour interventions^30–32^. Smartphone apps and wrist-worn activity trackers have increased physical activity in individuals with SCI, but these interventions did not target or measure sedentary behaviour^33,34^. Therefore, previous interventions may not target appropriate BCTs to effectively reduce sedentary behaviour in individuals with SCI^8^. The technology-based component in the current study has been co-designed with a specific focus on sedentary behaviour, which is likely to lead to an acceptable and effective intervention.

Education is included in the present intervention, in line with an intervention co-produced with stroke survivors using the BCW^20^. This highlights the importance of education around sedentary behaviour in individuals with neurological conditions^20^. A review found that interventions implementing education were effective for increasing leisure-time physical activity in this population^35^, however effects on sedentary behaviour have not been examined. Interventions utilising education have shown promise for reducing sedentary behaviour in non-disabled individuals^36,37^. The present study provides novel findings that an intervention targeting reductions and breaks in sedentary behaviour should include an educational component.

Goal setting is an important intervention component for targeting sedentary behaviour in individuals with neurological conditions, as found in a co-produced intervention in stroke survivors^20^. Reviews have demonstrated the effectiveness of goal setting for reducing sedentary behaviour in non-disabled individuals in workplace^38^ and non-workplace settings^36^, hence it being likely that goal setting will be effective in the present intervention. Motivational support was included in the present study to aid goal setting and working towards goals. The inclusion of motivational support is backed by a meta-analysis demonstrating that motivational counselling is effective for reducing sedentary behaviour in clinical populations^30^. In line with findings regarding facilitators for physical activity in individuals with SCI^39^, peer support was also considered a potential strategy for reducing sedentary behaviour in the present study. Therefore, motivational support and peer support may both be effective for reducing sedentary behaviour in individuals with paraplegia.

Consistent with physical activity research, providing activity tools was identified to help overcome barriers related to affordability and access in individuals with SCI^39^. Adding objects to the physical environment, such as wearables devices, sit-stand desks and exercise equipment, is an effective intervention strategy to reduce sedentary behaviour in non-disabled individuals^36,37,40^. However, there is limited evidence evaluating the use of activity tools to enable regular ‘activity breaks’ throughout the day. The inclusion of activity tools is, therefore, a novel component for reducing sedentary behaviour.

Some participants with paraplegia raised concerns with the term “sedentary behaviour” due to the stigma attached around wheelchair use. Instead, these participants suggested referring to “inactivity”, in line with findings in stroke survivors^20^. This suggests that alternative terminology and/or definitions relating to sedentary behaviour should be developed for individuals with physical disabilities. Some participants with paraplegia also preferred *positive* messaging in relation to the target behaviour (e.g. doing *more* activity, instead of engaging in *less* sedentary behaviour), as was also expressed by stroke survivors^20^.

### Strengths and limitations

Strengths of this study include the use of co-design combined with the BCW to undertake a rigorous and systematic approach to intervention development. The intervention is generalisable to a diverse range of individuals with paraplegia due to the varied sample of participants at different stages of the SCI healthcare pathway, inclusion of multiple key stakeholders and PPI.

Limitations of the study include the sample size being smaller than planned and comprising only Caucasian participants. A representative view from these participants may, therefore, have not been achieved, potentially limiting generalisability of the intervention. Not all participants were available to participate in both workshops, which is inconsistent with a traditional co-design approach. However, this approach provided the opportunity to understand acceptability of intervention options from individuals who were not involved with developing the initial concepts.

## Conclusion

This study reports on the development of a novel intervention targeting sedentary behaviour in individuals with paraplegia. The intervention was co-designed using the BCW, with input from multiple stakeholders, meaning that the developed intervention is likely to be feasible, acceptable and effective for end-users. It is recommended that future interventions are developed utilising a combined approach of participatory methodology and behaviour change theory to optimise acceptability and potential effectiveness. The feasibility, acceptability and effectiveness of the intervention is being evaluated in a further study. This research could inform public health and clinical care guidelines with a focus on sedentary behaviour in individuals with paraplegia.

## Data Availability

Excerpts of anonymised quotes included in the framework analysis are available in Supplementary Table 1.

