## Supplementary Table 1 for "Using the Behaviour Change Wheel to co-design a sedentary behaviour intervention in individuals with spinal cord injury"

**Supplementary Table 1.** All quotes for barriers and facilitators for reducing and breaking up sedentary behaviour

| COM-B construct | COM-B micro-construct | TDF domain | Barrier or facilitator identified (n = number of participants that identified it) | Illustrative quotes |
|---|---|---|---|---|
| Capability | Physical capability | Skills | Fatigue (barrier) (n = 10) | HCP (Physiotherapist, Male): "So I suppose my other thoughts is that rather than challenges more from from awareness, is that because they're using their shoulders much more often, is that we teach and advise around shoulder protection, and so therefore, if you're breaking up the sedentary behaviour actually an important part of shoulder protection is rest. And so I guess in the chair for a long bit of time, wheeling around a lot, transfers, everything like that. You're using it a lot more that it is actually designed to do" .<br><br>HCP (Physiotherapist, Female): "And I guess we've talked a lot about wheelchair users, but we know that the majority of our patients come to us incomplete. So most of our patients will spend some time on their feet, umm. And. But equally, most of our patients will at best use a combination of a wheelchair and ambulation to get around. And again, the physical effort that's involved in getting up and moving about on your feet is prohibitive for lots of people".<br><br>CCG (Charity worker, male): "When you're newly injured, predominantly a lot of your time is spent on personal care and just getting the day to day done. That can be hugely, hugely physically demanding as well as psychologically".<br><br>PwP (T4 Complete, Male): "I think that's similar to what I do. In my workshop, I restore things, up cycle things. Yeah. And so and also my wife gets to be doing the DIY jobs which, it's amazing what she'll let me do when it's a DIY thing, but if I wanted to do something, then "oh no, no, no, that's far too |

| | | | | |
|---|---|---|---|---|
| | | | | dangerous. No, you can't be doing that". And but I go all day. And by the evening, I'm knackered".<br><br>PwP (T4 complete, male): "The other spinal cord patients I've talked to, people tend to get tired more quickly than you would be if you were not in a wheelchair".<br><br>PwP (T12 Incomplete, Male): "I'm deliberately trying not to cram too much in, as I know it makes me tired".<br><br>PwP (T4 Complete, Male): "So I know I say it again and again, but today will be tiring for me. I was up an hour earlier than I normally am, cab journey here for 45 minutes, doing this, travelling home. I will be tired, so knowing that I was doing something equally kind of time consuming on Wednesday, I've left tomorrow to be a down day".<br><br>PwP (T12 Incomplete, Male): "Yesterday was marked as a down day because I've been doing today. Because sitting up too long, your feet swell up, then I start throbbing like he said. Yeah, you can't feel it, but they swell up massive. You gotta put them above your head, on the bed. Look, I've got a hoist thing. That's where I put them, I lift them up and I leave them. Just hanging there all night just to drain the fluid back into my body. And it's the pain, like if I'd have done loads yesterday, I wouldn't be here today. Even driving in my van, I start getting pain. Then I start getting pain and my shoulders start and it spasms your back. Like I said to you, didn't I? My sitting down is your standing up all day".<br><br>PwP (L1 Incomplete, Male): "And I think that's the thing with a wheelchair, though, isn't it? So, like my mother said, I haven't sat down all day. And I go well I've sat down most of the day, but I'm knackered. You know, and it's, if I know I'm sort of |

| | | | | |
|---|---|---|---|---|
| | | | | relaxed, if I transfer to the sofa and it's evening time watching TV, or just had dinner, or having dinner or whatever".<br><br>PwP (L1 Incomplete, Female): "I find getting in out the car tiring cause of lifting my chair in".<br><br>PwP (T11 Incomplete, Female): "Yeah. So that was me. Just generally, you know the increased level of fatigue maybe just because I'm newly injured. But that sometimes makes it difficult. Sort of 4:00 PM for me as a cut off point to become active throughout my day. But then I'm up at 5:30, so I think it's just generally what your routine, you know, how that's it. So that is a new thing for me".<br><br>PwP (T11 Incomplete, Female): "Not really. I mean, I'm kind of all out though, until about half four. Then that's sort of my calming down time. I would probably do less transfers, a little bit sitting in front of the television. But then putting something up. But that's the time at which I've noticed the most drawback from for being active".<br><br>PwP (T4 Complete, Male): "Yeah, I mean, I'll get to sort of like the evening. And sometimes I'm actually fine and I'm bouncing around and some of the days we've been out and done a lot in that daytime. Yeah. You do feel tired quicker in a wheelchair than you did pre wheelchair life. Yeah. Do you know what I mean? I could do a lot more pre wheelchair life than I can now without feeling shattered. So yeah, fatigue is one. It is tiring being a wheelchair. And people don't seem to understand that. But it's not only physically tiring. It's mentally tiring because how you're having to flip things in your brain all day long? Have you … *inaudible* to be positive so you can do things? That gets meant. That does your head in towards the end of that and you are mentally shattered as well as physically shattered?". |

| | | | | |
|---|---|---|---|---|
| | | | | PwP (L1 Incomplete, Female): "Yeah, I agree. I I get tired quite easily. And I mean, I take a mid-afternoon nap 'cause. I'm awake at 4:00 in the morning. Take mid afternoon nap and then go to bed, sort of ten-ish. And I don't do much in the evening, I tend to sit and watch telly in the evenings, and I don't go out in the evening 'cause it's too much for me".<br><br>PwP (L1 Incomplete, Female): "I do nothing the day before, yeah. And the day after as well". |
| | | | Personal care routine (barrier) (n = 10) | HCP (Physiotherapist, Female) "There's less flex in their day, they've got less free time available if they're also a working person to fit that in because, yeah, they're, potentially some of them [have] got longer morning routines".<br><br>HCP (Physiotherapist, Female): "Yeah. Or in and out of the shower in 10 minutes. You know an hour would be a speedy morning routine for somebody who needs to manage their bladder and their bowel and get washed and get dressed".<br><br>CCG (Charity worker, Male): "When you're newly injured, predominantly a lot of your time is spent on personal care and just getting the day to day done. That can be hugely, hugely physically demanding as well as psychologically".<br><br>PwP (T12 Incomplete, Male): "Yeah, getting dressed, your. Yeah, cos when you get dressed you're flipping all over the bed".<br><br>HCP (Physiotherapist, Female): "We practice sort of out outward activities and it's it's RAs and RPs um who are more responsible for getting people out and about and having the experience of being in social environments away from the spinal cord injury centre. Other things that are really challenging is managing bowel routines, I mean bladder routines. So if people use intermittent catheterisation, they have to do that regularly. |

| | | | | |
|---|---|---|---|---|
| | | | | And that's not always easy in every environment, and you know, some people might need to do a catheter every four hours. People have to monitor how much they drink. So you you can't drink freely necessarily, if that's, if that's part of your caring team. So again, all of that alters how spontaneous you can be".<br><br>PwP (T12 Incomplete, Male): "It's catheterisation. Do you catheterise? Self-catheterise? You start getting bladder infection, and then start sweating. Then most people get tremor relaxed because they get a bladder infection. Mine increases pain. If I was sat here now and my knee was resting against that, I'd know about it. Even though I can't feel it, something in my head like is telling me something is not right".<br><br>HCP (Physiotherapy Student): "I think also because quite a lot of the patients, because the transfer can be quite, like a lot of energy is quite challenging. But quite they'll want to prioritise what transports they're doing during the day. For when they need them in function, so trying to reduce unnecessary transfers where you can".<br><br>PwP (T6 Complete, Male): "Whereas myself, I have to get a banana board, slide myself into the car. And then start dismantling the chair. I'm putting it in, but yeah. By the time you've done that, sometimes you have to sit for 5 minutes before I start. I'm knackered".<br><br>PwP (T9 Complete, Female): "Transferring to my shower chair, and then wheeling into the bathroom. So, I've got a double kind of transfer".<br><br>PwP (T6 Complete, Male): "That's just in the car. So if I was going, I mean this is gonna sound stupid. If I'm at home and I transfer on to the toilet, when I, once you come to where you're dressing yourself again, I can't redress myself, right? So I then |

| | | | | |
|---|---|---|---|---|
| | | | | have to get onto the floor. Shuffle. Bum shuffle, going back to what you were saying earlier. Roll around, like a kind of beached whale, and then into my hoist, we start back into my chair again".<br><br>PwP (T4 Complete, Male): "Uh, no, so for me, to Doug's point, to Christine's point, so I can transfer from bed to commode or bed to chair umm, and from commode to bed. and bed to chair, and chair to bed quite quickly. Yeah, just. Yeah. Couple of minutes, no more than that umm. But then, you know, I have to, once I have a shower, I have to transfer back to bed, to get finished getting dressed, getting dressed, then sit up again. For me, it's the going from lying to sitting in bed position. It's hard. Hardest. I have help. I've got, you know, you know, getting better at it. Umm. So in terms of kind of, you know, going for a wee, you know I don't get out my chair. I don't. I don't. Yeah, I won't go to toilet here and transfer. Yeah, I will just stay sitting in my chair".<br><br>PwP (T11 Incomplete, Female): "I used to go the gym before my accident. I think the barrier for me mainly is the amount of transfers that you would have to do. You know, I'm still trying to stay as active as possible, but I think. You know the barriers behind transferring, driving, whether the gyms are accessible or not. I think it just makes it an extra layer to have to contend with".<br><br>PwP (T11 Incomplete, Female): "Yeah. So I am, you know, with a spinal cord injury, you have. I have to use catheters to go to empty my bladder. So I will deliberately get up, go over to the toilet rather than using bags. That's something that we were that. That's a way for me to be able to kind of move or make me make myself a cup of tea. I might get up out of the chair to do that, but I do a lot of transfers so I don't stay in my chair very long. I will transfer out of my wheelchair onto the sofa or, and |

| | | | | |
|---|---|---|---|---|
| | | | | out to the kitchen. And so I think I just. I don't sit still long enough. I also have five children so I can't sit still for very long periods of time”.<br><br>PwP (T4 Complete, Male): “Well. Yes, it's the on on the on. I’m the same as you that I'm never in my wheelchair. If I've got an option to be out my wheelchair, even in a coffee shop, I'll jump into a tub chair, I'll jump into a sofa in the coffee shop, my sofa at home. I just. My wheelchair’s a pain in the neck of the morning and I hate it. Yeah. So if I if I can limit my time in there to the best I possibly can, I will. Whether I'm sitting anywhere but in my wheelchair. Transfer to the car, go down the shops, do this, do that. Yeah. I'm just bouncing around all day”. |
| | | | Pain (barrier) (n = 7) | HCP (Physiotherapist, Male): “So I suppose my other thoughts is that rather than challenges more from from awareness, is that because they're using their shoulders much more often, is that we teach and advise around shoulder protection, and so therefore, if you're breaking up the sedentary behavior actually an important part of shoulder protection is rest. And so I guess in the chair for a long bit of time, wheeling around a lot, transfers, everything like that. You're using it a lot more that it is actually designed to do”.<br><br>HCP (Physiotherapist, Male): “I suppose our mindset probably goes a lot towards around how they strategically used their their energy in the day, from a pace perspective. We kind of frame sedentary behaviour as that sort of rest period. As opposed to, yeah, breaking it up per se. So when you kind of frame it in that way in order to mitigate these secondary complications such as shoulder pain et cetera”.<br><br>CCG (Charity worker, Male): “I I I think the pain is such a personalised thing, some somebody just because of your level of. I mean, if you've got neurological pain or shoulder pain or you know, age-related pain, then that's going, that's going to |

| | | | | |
|---|---|---|---|---|
| | | | | influence it. Yeah, so so pain. So pain. So pain is a is a personal thing really".<br><br>PwP (T6 complete, male): "I would say the only thing that's increased my sedentary behaviour is pain".<br><br>PwP (L1 Incomplete, Male): "I'm not doing anything, I'm just sitting there because I have to have my legs straight to stretch my leg, and then it's my legs".<br><br>PwP (L1 Incomplete, Male): "I don't get pain just discomfort. OK, discomfort, I thought. Yes, for some people might say it's like always like. So it's like between nought and 10, but some people go off high level 2, but I've got quite a high pain tolerance. So for me it's like "oh it's uncomfortable"".<br><br>PwP (T12 Incomplete, Male): "Yeah, because I can't do anything when that's. When that's when my knee starts".<br><br>PwP (T6 Complete, Male): "Pain is the main one [that affects my sedentary behaviour]".<br><br>PwP (T12 Incomplete, Male): "Pain will stop me doing anything. Wheelchair skills won't because I'm pretty good in a wheelchair".<br><br>PwP (T1 Incomplete, Male): "Yesterday, for the last week I've been in agony and I just couldn't bother and I just didn't".<br><br>PwP (T4 Complete, Male): "If I can just say one more thing, one more thing very, very quickly, if that's OK. Is that about it is obviously the pain factor. Yeah, but I find that if I train every day, day in, day out. For the first 5 minutes, I'm not lying, it bloody hurts. Yeah, but once you get, but once you get a bit of warm, warm blood flowing around the muscles. Yeah. And |

| | | | | |
|---|---|---|---|---|
| | | | | warm oxygen. Yeah. It doesn't hurt anymore. Yeah. So it's getting over that. Yes, it's really hurts. This is painful. Yeah".<br><br>PwP (T4 complete, male): "If you sit still and don't move, then when you do move, it bloody hurts. Simple. You sit there and don't do nothing [anything] all day". |
| | | | Comorbidities (barrier) (n = 4) | HCP (Physiotherapist, female): "People have comorbidities and it's an aging population. And so you run into all the same problems with physical activity you do with anybody else".<br><br>CCG (Charity worker, male): "There might be, you know, underlying health issues that are going on or which might be spinal or non- spinal… your general health can have quite a big impact on your ability to be, you know, sedentary".<br><br>PwP (T6 Complete, Male): "Also it's the drugs [that I take for comorbidities that affect sedentary behaviour]".<br><br>PwP (T1 Incomplete, Male): "Well. I I laugh because I was just like PwP until three years ago when I went in for an operation in hospital. They dumped me at home on my own. I got sepsis. And you can see the size of me. But I've, I mean, you know, the sepsis down here ruined me, and it knocked all all the feeling out of me. I couldn't do anything". |
| | | | Injury (barrier) (n = 4) | HCP (Physiotherapist, Male): "Get a shoulder injury as a wheelchair user, it's so it's so hard to reverse, yeah".<br><br>PwP (L1 incomplete, male): "that happens quite frequently, you pick up injuries or stuff happens, you know, that knocks you back".<br><br>PwP (T9 Complete, Female): "So, yeah, just a broken leg, for instance, that would lay you out for six weeks, wouldn't it". |

| | | | | |
|---|---|---|---|---|
| | | | | PWP (T12 incomplete, male): “When I broke my leg, that's six months in bed”. |
|  |  |  | Pressure ulcers (barrier) (n = 4) | PwP (L1 incomplete, male): “You know, if you’re on bed rest for six months... you know, you had a sore or something”.<br><br>PwP (T12 Incomplete, Male): “It went right up into the bone. And it went into pelvis. My leg just swelled up. They thought it to cyst, so I went to the first hospital. They cut it open left the wound open. Six months in bed, still a wound open, weren’t healing up”.<br><br>PWP (T4 complete, male): “You could be bedridden for long periods of time [if you got a pressure sore]. We heal a lots more slowly than able-bodied people”.<br><br>PwP (T12 Incomplete, Male): “Oh if I got pressure sore and I’m in bed, it annoys me. Yeah, because I'm in bed for at least six weeks”.<br><br>PwP (T1 Incomplete, Male): “I got a taphylococcus Auris infection and I was just stuck on my stuck on my bed. Almost immobile for six weeks and that gave me a bed sore”. |
| | | | Lack of physical function (barrier) (n = 4) | HCP (Physiotherapist, Female): “They can only use their arms to do it [break up sedentary behaviour with physical activity]”.<br><br>CCG (charity worker, male): “The higher your level of injury, then obviously the lower the amount of muscle groups that you've got to use”.<br><br>CCG (Charity worker, Male): “That's going to then influence things like you know how high you can get your heart rate. It's going to, it's going to make you think, well, I don't have, you know, if you're tetraplegic. Well, I don't have much use of my arm. So how am I going to… how is that going to benefit me? |

| | | | | |
|---|---|---|---|---|
| | | | | Whereas if you're a lower level of injury then you're potentially going to be able to do a lot more more exercises. But I don't. So does that make sense?”.<br><br>PwP (T4 complete, male): “A T12, she will have core muscles and be able to put both arms out in front of her… Whereas me, I'm at 5. If I do that, I'll fall out [of] my wheelchair”.<br><br>PwP (T1 Incomplete, Male): “So there's two aspects for my exercise thing. I mean, I don't know you can decide, but one's the mental drive to do it and some of us have had it kicked out of us. The other one is the physical ability to do stuff. And then that's it really. Pop that in your things to do”.<br><br>PwP (T4 Complete, Male): “I think I think the biggest thing is is. The biggest thing about doing anything is that you've just gotta just you just gotta be able to accept that it’s bloody frustrating. Yeah. What used to take you two minutes to do or five minutes to do, might take you 15 minutes. Yeah. Silly little things like cutting things up. Yeah. Obviously. Because I'm a T5, I've got no tummy muscles, so I can't use both hands”. |
| | | | Transfers (barrier) (n = 4) | HCP (Physiotherapy Student): “the transfer can be quite, like a lot of energy; it’s quite challenging. But quite they’ll want to prioritise what transfers they're doing during the day, for when they need them in function. So, trying to reduce unnecessary transfers”.<br><br>HCP (Physiotherapist, female): “Transfers are more difficult. All of these things, because the first chair that they get from their wheelchair service is rubbish”.<br><br>PwP (T11 incomplete, female) “I used to go the gym before my accident. I think the barrier for me mainly is the amount of transfers that you would have to do”. |

| | | | | |
|---|---|---|---|---|
| | | | | PwP (T9 Complete, Female): ”Transferring to my shower chair and then wheeling into the bathroom. So, I've got a double kind of transfer”. |
| | Psychological capability | Knowledge | Lack of knowledge around sedentary behaviour (barrier) (n = 15) | HCP (Physiotherapist, Female): “They might be quite offended to say that sedentary behaviour is sitting down and breaking in sedentary behaviour is standing up”.<br><br>HCP (Physiotherapist, female): “There isn't specific guidance on sedentary behaviour. So there, if there isn't specific guidance, it's hard for people to have knowledge”.<br><br>HCP (Physiotherapist, Female): “And like I said, I think. Like we don't know how much activity in a paraplegic is the difference between being unhealthily inactive and healthily inactive. It’s very hard to make people. Um. Give good education on that front when we don't actually have an understanding of what what good looks like. So from a professional’s point of view, that's a challenge. But yeah. Go on”.<br><br>HCP (Physiotherapist, Female): “Yeah, because I don't. I think people who haven't come through a spinal cord injury centre, I'm not sure they would have even the advice we give about the exercise recommendations. Certainly when I was I was working, going out to visit people in hospital when they were first injured, that is far from this sort of stuff we're discussing, it's it's much too early. And then they go somewhere else and it maybe never gets mentioned if they don't come through a spinal cord injury centre. So there may be people out there with very different knowledge depending on their their rehab pathway”.<br><br>HCP (Physiotherapist, Female): “I also don't know if we speak about breaking up sedentary behaviour particularly well in all honesty. Thinking about it, we talk about exercise all the time and we talk about shoulder care and preservation of shoulders and we talk about transfers and pacing and exercise and going |

| | | | | |
|---|---|---|---|---|
| | | | | to the gym, going swimming, going out and pushing and we talk about all that. And I think we do that pretty well. But just considering what we're talking about in terms of breaking sedentary behaviour, I'm not 100% sure I actually go through that with patients".<br><br>HCP (Physiotherapist, Female): "I'm quite sure that I don't, but what I think is that asking someone to push across the room is insignificant. If you sit still and watch the telly and then you push across the room and come back again".<br><br>HCP (Physiotherapist, Female): "Yeah, completely. But I wouldn't advise people on moving about more because I I haven't got any understanding of if that is in any way worth their time and thought. And and my guess is perhaps it's really not".<br><br>HCP (Physiotherapist, Male): "Something for you guys, because I think it's it's really challenging is that when when you, when you problem solve it around what do we define as sedentary behavior for these different kinds of people".<br><br>CCG (Charity worker, Male): "That we forget the fact that any movement is good movement. Any activity is is good activity. So it's it's trying to make people aware… So I think it's just helping people to understand what do we mean by you're talking about 5 minutes of exercise. People really need to be made aware of what that looks like, what that could be, and also and also what the benefits physiologically and psychologically are going to be".<br><br>CCG (Charity worker, Male): "Yeah. Similar to what CCG was saying that some people can feel a bit intimidated by, you know, just looking at a sport like wheelchair rugby for example, that that any kind of, you know, immediately you're you think of those sort of high end or those elite sports whereas you know I |

| | | | | |
|---|---|---|---|---|
| | | | | sometimes try and say to someone that, you know, exercise can be anything, it can be just walking your dog. you know, just just getting out of the house”.<br><br>CCG (Family member, Female): “I think for my dad, it would be having the understanding that doing anything really would benefit him. Not in terms of sort of what you would sort of call, I guess he is not really capable of doing exercise or sport or such, but just for him to understand that to be doing anything really would be beneficial”.<br><br>CCG (Charity worker, Male): “There's also an element of. If you look at sort of like what positive psychology is all about, it's it's almost recognising that, the fact that you're, you know, that you're out of bed and that you're moving around already. That's a positive starting point that you've got. So just going through your personal care every day is, you know, or you know, keeping the house tidy and all of those things, that those are all contributing to what you're doing. So, for so many people, they're already being active. We're asking them to become more active. So I think it's sort of like helping people to understand that you're already doing something. We're just kind of like wanting to help you become more aware of how much, how active you are. And then also thinking about how we can, you know, do, do how, how could we introduce more activity to improve your situation”.<br><br>PwP (T12 Incomplete, Male): “Sedentary? Like laziness?”.<br><br>PwP (L1 Incomplete, Male): “I'm paralyzed from here down. So yeah, a good 80% of our body doesn’t work. Umm yeah, so I mean, am I sedentary now, sitting around doing this or or, not? And I think that's quite a key”. |

| | | | | |
|---|---|---|---|---|
| | | | | PwP (L1 Incomplete, Male): “[What is] the definition of being sedentary?”.<br><br>PwP (T9 Complete, Female): “Yeah, but, but we don't see that as exercise, do we? It's just getting from A to B. But it is exercise”.<br><br>PwP (T12 Incomplete, Male): “No they don’t give you [guidance on sedentary behaviour]”.<br><br>PwP (T12 Incomplete, Male): “There’s nothing regimental, no. They just try and teach you in hospital in order to send you out”.<br><br>PwP (T6 Complete, Male): “I’m going to say no as well. The only time they say [to reduce sedentary behaviour] is for to stop you getting sores”.<br><br>PwP (T6 Complete, Male): “That that's the big word, sores. And apart from that you don't get told about movement at all”.<br><br>PWP (T4 complete, male): “I don’t think the word sedentary was ever said [during in-patient rehabilitation]”.<br><br>PwP (L1 Incomplete, Male): “Because isn’t it, what they say about something in motion stays in motion, rather than something stopped. But it's more sort of difficult. Maybe it's in branding. Whatever it is, what you're trying to achieve in a way where it, because people are right, you're right. People are like “oh exercise, God that makes me sick”, you know?”.<br><br>PwP (T1 Incomplete, Male): “OK. Could you remove? I'd be cautious about using if you're gonna spread this out. The word sedentary means you sit on your bum all day. We don't have a choice. So I would say “inactivity”, you know? Yeah. Be cautious. I I'm not offended by that. I'm used to being called a |

| | | | | |
|---|---|---|---|---|
| | | | | cripple. And I I know it. I don't mind. But some people get offended by someone saying you're, you know, sedentary, especially if you haven't got a choice. So I would take the word out".<br><br>HCP (Physiotherapist, Female): "They might not wanna look at it. But, it's just there. Is it another option, potentially? Rather than just going 'be active, you need to move' and people go 'don't want to'. Actually go 'OK, you're not moving, lets maybe think about why you are not moving, and actually this might be a good opportunity to link in with this person or talk to this person or watch this podcast or these are different options potentially that you use'. Instead of 'move now'".<br><br>HCP (Physiotherapy Student): "You just absolutely have to move someone from my extrinsic to intrinsic motivation. And that might start with having like having therapy sessions and then even like within those sessions, just discussing like, why you would want to continue these after. But I'm sure that's already done".<br><br>CCG (Charity worker, Male): "Yeah. Similar to what Will was saying that some people can feel a bit intimidated by, you know, just looking at a sport like wheelchair rugby for example, that that any kind of, you know, immediately you're you think of those sort of high end or those those elite sports whereas you know I sometimes try and say to someone that, you know, exercise can be anything, it can be just walking your dog. you know, just just getting out of the house".<br><br>CCG (Family member, Female): "I think. I think it would be helpful. In the case of my dad, really, for my mum to understand that it's good for him to actually be doing as much as he can. I think as long as a family member are aware of bits of it". |

| | | | | |
|---|---|---|---|---|
| | | | | PwP (T1 Incomplete, Male):<br>Or, you know, and then we get through the incentivizes, you get the lies for parents. So, the devil will come and get you if you pick your nose, you know, tonight, you know, whatever, you know what I mean. But you need to make me more positive. But you can do that. You can. That's one aspect from, you know your your study if that's a possible thing. You can say “reasons why not”, “reasons why” and then if you can't win the people over then they go back to the the the, you know, the what do you call it, intellectual aspects rather than calling them mental, the intellectual, and then you got the physical”.<br><br>PwP (T4 Complete, Male): “For me it was a bit of both really. I enjoyed standing up and having a conversation with someone at eye line, yeah, rather than looking up or looking, them looking down on me. And also, you know, I mean when you're standing up, yeah, it's good. It affects your blood pressure, your heart bounces more. Yeah. Do you know what I mean? To pump the blood around your body more. So I think gravity, it helps your lungs go down, digestive, and stuff like that. So I think standing is really important”. |
| | | | Knowledge of pressure relief (facilitator) (n = 6) | CCG (Charity worker, male): “Something that all wheelchair users are told to do is a huge part of their education when they're in hospital is to do pressure relief to make sure every hour or so that they make sure they shift in their chair and lean forward in order to avoid pressure sores”.<br><br>PWP (T4 complete, male): “Part of my routine would then be well, I need to do pressure relief… And therefore, you know, that's exertion”.<br><br>PwP (T4 Complete, Male): “So they they recommend that every two hours sitting in your chair that you do 2 minutes of [pressure relief activities]”. |

| | | | | |
|---|---|---|---|---|
| | | | | PwP (T12 Incomplete, Male): "They used to tell me every 20 minutes you've got to lift up just to get some blood on your bum. Otherwise you can't feel your bum. But my sitting down is not standing up. My lying down is your sitting down".<br><br>PwP (T4 Complete, Male): "But let's just stay in the leaning forward. Yeah, and yeah, parts of your body which will hit the chair. to be not touching the chair".<br><br>PwP (T12 Incomplete, Male): "I got told [to do pressure relief] every 20 minutes".<br><br>PwP (L1 Incomplete, Male): "I was gonna say from what they advised you, I'm sure it's every half an hour. They said to something on those those lines".<br><br>PwP (T12 Incomplete, Male): "Even just moving in your Chair, sitting forward like that. Yeah, that's pressure relieving".<br><br>PwP (T9 Complete, Female): "Yes, so I thought "this will clear up, this will clear up". But actually it didn't, so I had an operation and… Two years later, I managed to break my leg. And so it was sort of, you know, and it was full cast umm, so and I you know, I was still using the shower chair, so I put extra weight on back side. So I had to have a second operation. But yeah, just leaning forward makes a difference. So you don't physically have to lift or just won't be normal side or untouched. Good".<br><br>PwP (T1 Incomplete, Male): "One other thing for you [the researchers] is the other benefit about doing transfers, get moving, is you don't get bed sores". |
| | | Memory, attention, and | Reminders to be active (facilitator) (n = 5) | HCP (Physiotherapist, Female): "What about like an app that beeps at them, it says 'move now'". |

| | | | | |
|---|---|---|---|---|
| | | decision processes | | CCG (Family member, Female): “Can I also add something if that's alright? So there was there was a bit of an observation from my part when my sister had her accident. She actually gave away her apple. She used to have an Apple Watch. She gave away her Apple Watch because it kept notifying her to move to stand up. So there's a little thing on the watch. It’s kind of a push, a push notification that says “stand up” and tries to keep you kind of moving. And she found that really triggering, but it's almost it's almost that kind of useful tool perhaps of, you know, encouraging somebody to move rather than rather than, you know, stand up as the term. But I think she would really benefit from almost, like, a smart device that is inclusive for those that are wheelchair users to be able to have that inclusivity with everybody, she would, I think she would really benefit from something similar to that”.<br><br>CCG (Family member, female): “Definitely that thing of having some kind of prompt I think would definitely be helpful for him [father with paraplegia]”.<br><br>PwP (T12 Incomplete, Male): “No, you need it. You need your bum kicking sometimes”.<br><br>PwP (T12 Incomplete, Male): “The text message, everyone goes “read it”. You’re having a chat and go “oh, my phone”. With the big big “get up. Do something”. Yeah, it works. It will work”.<br><br>PwP (L1 Incomplete, Male): “I mean you've you've basically, for having the app and signing up for it, you're accepting, right? That it's gonna beep at me, it's gonna nag me. But I want that to happen because I know sometimes I can be a bit lazy or or ignore what I should be doing, you know? And it's interesting what you were saying about the sort of misconstruing between being sedentary and exercise. You know it's more. It's not so |

| | | | | |
|---|---|---|---|---|
| | | | | much exercise, it's keeping in motion, isn't it? Yeah, because isn't it?".<br><br>PwP (T12 incomplete, male): "The prompts, you know, to the alerts to say. "get moving, get moving"… alerts to say, "come on, keep going. You're nearly there", so sort of motivating alerts [would help me be less sedentary]". |
| | | | Do not think about sedentary behaviour (barrier) (n = 4) | CCG (Charity worker, male): "It's not really the first thing that's on their mind, you know, they've got other issues to think about".<br><br>PwP (T12 Incomplete, Male): "I don't [think about sedentary behaviour]. I just get on my bed [at the end of the day]".<br><br>PwP (T6 complete, male): "I don't think about it [sedentary behaviour] during the day".<br><br>PwP (T9 Complete, Female): "Yeah, I mean, some days I don't, yeah, I'm like "I haven't left the house today", but then I know I'm going somewhere tomorrow. So like you know, I don't, I don't let it bother me". |
| | | Behavioural regulation | Getting feedback on progress (facilitator) (n = 6) | HCP (Physiotherapist, Female): "Honestly, I think the people for whom it speaks to it would reinforce what they're doing anyway and maybe help maintain their their motivation. You know, Strava, whatever, people who love it use that as a way of communicating about things that they enjoy and that does keep their fire burning. But for the people for whom it's not interesting".<br><br>CCG (Family member, Female): "My sister [with paraplegia] used to have an Apple Watch. She gave away her Apple Watch because it kept notifying her to move to stand up. So there's a little thing on the watch. It's kind of a push, a push notification that says "stand up" and tries to keep you kind of moving. And she found that really triggering, but it's almost it's almost that |

| | | | | |
|---|---|---|---|---|
| | | | | kind of useful tool perhaps of, you know, encouraging somebody to move rather than rather than, you know, stand up as the term. But I think she would really benefit from almost, like, a smart device that is inclusive for those that are wheelchair users to be able to have that inclusivity with everybody, she would, I think she would really benefit from something similar to that".<br><br>CCG (Charity worker, Male): "I use a, I use a. I've got a, I've got an app which is called "High intensity interval training app". And I just I set that up and I've got sort of like, you know, 5 minutes of exercise, 8 minutes of exercise, 10 minutes of exercise. And it just does 30 seconds on 30 seconds off for like 5 reps, 8 reps, 10 reps and and that just gives me like a little calendar with a little thing that bounces up. And so if I'm doing some activity, I'll I quite like that because then it shows you like a month. It shows you for like the month of September. It's like little shapes throughout the calendar that show me I'm being active".<br><br>CCG (Charity worker, Male): "There's also an element of. If you look at sort of like what positive psychology is all about, it's it's almost recognising that, the fact that you're, you know, that you're out of bed and that you're moving around already. That's a positive starting point that you've got. So just going through your personal care every day is, you know, or you know, keeping the house tidy and all of those things, that those are all contributing to what you're doing. So, for so many people, they're already being active. We're asking them to become more active. So I think it's sort of like helping people to understand that you're already doing something. We're just kind of like wanting to help you become more aware of how much, how active you are. And then also thinking about how we can, you know, do, do how, how could we introduce more activity to improve your situation". |

| | | | | |
|---|---|---|---|---|
| | | | | PwP (L1 incomplete, male): “It’d be good to like, keep a log of say, whatever activities you're doing. And having like your personal best”.<br><br>PwP (L1 Incomplete, Male): “Umm, well the lack of results one, goals. It could go back to the prompts, you know, to the alerts to say. “get moving, get moving”. I think it's been said before, alerts to say, “come on, keep going. You’re nearly there”, so sort of motivating alerts”.<br><br>PwP (T6 complete, male): “You can get an app for your phone, so you can actually see what [activities] you've done”.<br><br>PwP (T4 Complete, Male): “And yeah, I would say that at first, someone might need a need a high level of intervention. Yeah, but then that intervention might become less and less, which it should do as their mind becomes more down to them”. |
| | | | Creating schedules around activity (facilitator) (n = 6) | HCP (Physiotherapist, Male): “Strategic breaks of perhaps breaking up certain behaviours, yeah”.<br><br>HCP (Physiotherapist, Female): “There's another thing that happens is they can have quite a routine in the morning, can take a long time, so they got to do like getting washed getting dressed, but a bladder and bowel routine that takes them often longer in the morning to get out of the door than it would take us to get out of the door to work. So if they’re somebody who works, there’s less flex in their day, they've got less free time available if they're also a working person to fit that in because, yeah, they're, potentially some of them got longer morning routines”.<br><br>HCP (Physiotherapist, Female): “Lots of lots of our advice will be around people making choices about how they expend their energy. And there's the difference between a blast of activity versus a more continual activity across the day, and the |

| | | | | |
|---|---|---|---|---|
| | | | | morning routine is really physically taxing. So that's harder work for them than it would be for us, but we might advise people to use carers to assist them with that in order to save their energy for other parts of the day. And so there's a difference between the energy in the way that we think of, like your ability to get up and go and do things versus your metabolic energy cost, which for somebody who's a wheelchair user, will have bigger peaks and troughs. But you might imagine across the day is is less. But that doesn't make it any less tiring".<br><br>HCP (Physiotherapist, Male): "What we used to kind of do is like, practice discharges, kind of do like a Monday to Sunday timetable of their life and be like 'Right, what happens in the morning for you?'. Build a 24 hour sort of plan here and get sort of as close as we can prior to discharge as to what looks optimal from a pacing perspective and so, therefore, part of the goals in say, when I used to do community referrals, was tweak their 24 hours sort of routine. To optimize it in a way where they can do a tailored exercise program alongside the rest breaks in between… Perhaps something like that again, it's really hard. As part of the reintegration phase, when patients are here and they go home, could be something to template and then tweak when they're at home, when they actually go about their daily life".<br><br>CCG (Charity worker, male): "Trying to get a bit of a timetable even into your normal daily living and to and to incorporate the activity in into that".<br><br>CCG (Charity worker, Male): "There's something around timetabling. It's something about thinking about, again as well we're saying your personal care, so I know exactly what time I need to go up in the morning to ensure that whatever happens, I'll get out of the door at a certain time. So, these are what must be going through most people's heads when, you know, when |

| | | | | |
|---|---|---|---|---|
| | | | | they get up in the in the morning and have to do these things that are very different now. And and time is is is kind of key key to that”.<br><br>PwP (L1 incomplete, male): “Yeah, prioritising your schedule to accommodate [for physical activity and sedentary behaviour”.<br><br>PwP (L1 Incomplete, Male): “So that's what I was gonna say when we're talking about the ones above. So like, time is looking at your day to day sort of calendar and what responsibilities you've got umm and the last one which”. |
| Opportunity | Physical opportunity | Environmental context and resources | Provision of information and/or opportunities for physical activity and exercise (facilitator) (n = 12) | HCP (Physiotherapist, female): “They'll sometimes tap into the charities as well. So they're going out and about and doing a bit more activity with some of the charities, so again, they'll start to overcome some of those barriers and see how they can plan things with the help of external sources”.<br><br>HCP (Physiotherapist, Female): “We always point them towards the charities, and the psychology team do an awful lot about pointing them towards their relevant charities and it is something we speak about. So, we do get some charities in to do some of the Backup wheelchair skills for us as well. So, they are signposted. But again, it's it's them taking that step forwards to actually tap into some of those support networks”.<br><br>HCP (Physiotherapist, Female): “They can also, Wheelpower is a charity that provides free Therabands, so any anyone who's called injured can ask for those. And they're like quite posh ones with handles on and stuff. They're they're good”.<br><br>HCP (Physiotherapist, Male): “Well, I was gonna say as well, who might be pretty well-placed in primary care services is Spinal Injuries Association, Back Up. they’re perfectly placed for |

| | | | | |
|---|---|---|---|---|
| | | | | that sort of thing [providing advice and resources related to activity]".<br><br>CCG (Charity worker, Male): "Mainly what we [Charity] do is as an information service".<br><br>CCG (Charity worker, male): "Wheelpower has probably got the best examples at the moment of showing they they've, they've got yoga, they've got people doing fitness classes and people doing different exercise activities".<br><br>PwP (T6 Complete, Male): "But, yeah. Where you are now, at Stanmore, there's gonna be loads of that type of thing, and then you can just pick and choose. That's why Stanmore's so good".<br><br>PwP (T4 Complete, Male): "Not particularly no, I'm not particularly sporty. Or, having said that I'm gonna go and do the inter-spinal unit games".<br><br>PwP (T6 Complete, Male): "Yeah, yeah. Like, you were saying earlier about, was it Sportability [that provided opportunities to do physical activity and/or exercise]?".<br><br>PwP (T12 Incomplete, Male): "Yeah, they do different areas, Sportability. Like you could run a group where go sailing. I could run a group to go shooting, you know, like target shooting".<br><br>PwP (T12 Incomplete, Male): "You could run a good doing archery in your area. I'm in [LOCATION REMOVED]. Yours is [LOCATION REMOVED]. That's what like, everyone organises it. Like, I would organise a group like this for about 8 people to go shooting.<br><br>PwP (L1 Incomplete, Male): "So going to financial, what you're saying about equipment, I got a small grant I think it was from |

| | | | | |
|---|---|---|---|---|
| | | | | Wheelpower or one of the charities. And so it was to enable me to go walking with my wife with the dogs.”.<br><br>PwP (T9 Complete, Female): “Well I’ve got, from Wheelpower, I’ve got some stretchy bands”.<br><br>PwP (T1 Incomplete, Male): “You get one that holds your neck up and it holds you and you can float and then you can move your fingers or your arms or your neck or whatever. Or anything else you can, but that's what and there's a social thing there where you can get lots of other people off of Physiotherapists or retired Physiotherapists who help you. And it's peanuts. It's about £5 where it would cost you £100 to do it in a normal pool”.<br><br>PwP (T1 Incomplete, Male): “People can go out and find stuff. And if you're prepared to go out the house, you could ask your doctor to recommend a Physiotherapist who will come to you. They do a six week. They used to do a six week course for the disabled, to rehabilitate them, and you get a membership with the gym for a huge discount”.<br><br>PwP (L1 Incomplete, Female): “Well, we used to have, locally, we used to have a scheme where we had volunteer befrienders. And I had someone who used to come once a week from that [giving her social opportunity to go out and be active]. But they stopped running the scheme”. |
| | | | Lack of wheelchair accessibility in and outside of the home (barrier) (n = 10) | HCP (Physiotherapist, Female): “Sometimes people can't get out of the house as well, and I think then it's the accessibility which I'm assuming you might move on to in a minute. The accessibility of things like even just going on a pavement is extraordinarily difficult for some of our patients as well. Yeah. Ramps. Stairs. steps. Getting in and about the house sometimes”. |

| | | | | |
|---|---|---|---|---|
| | | | | HCP (Physiotherapist, Female): "Some patients have got the higher wheelchair skills and ability, but I think it's also the setup of the environment as well and because it may be all great that they can get up downstairs potentially in a very um, sort of protected environment here potentially. But then we all know getting in and out of that house is very different because you've got to get over the the potentially the lip of the doorframe and you've got the stairs outside potentially, which may be very narrow. May be very difficult to navigate".<br><br>HCP (Physiotherapist, Female): "I guess being mobile in a peer group is also more difficult. So lots of us will go out with people who are similarly able, you know, for a walk or to a restaurant or a bar. And for lots of people who are wheelchair users, that becomes much less part of their life, but it doesn't have to be. And some people find a way to make that work, and it's fine. But for lots of people that falls out of their routine because their challenges in accessing environments are different to the rest of their peer group, and that is a barrier to them".<br><br>PwP (L1 Incomplete, Male): "Yeah, well, that mine came about, I think from shuffling on my backside as well because it was early after my injury and I was getting used to getting around and I think we were staying at mother-in-law's flat, which is upstairs. No space for a wheelchair, so I would stand and hold on to walls or furniture. But what was safe? My wife's words was, you know, "shuffle" and that. Yeah".<br><br>PwP (L1 Incomplete, Male): "It could be. I mean, we're lucky because we bought our house as we were coming back from the States and we gutted it and did what we were doing it before we moved in. And a lot in our mind was sort of open plan, or enough room for the wheelchair to work, you know. And and I had this conversation with mt wife actually just the other day and talking about going to visit other people in their houses |

| | | | | |
|---|---|---|---|---|
| | | | | and how difficult it is because there are thresholds, there are steps to get in. They probably don't have enough room to wheel a wheelchair around, cause some people's houses can be really old and they've got really sort of steep steps”.<br><br>PwP (T12 Incomplete, Male): “You can’t get in the bathroom and people’s kitchens”.<br><br>PwP (L1 Incomplete, Male): “Some of the bathrooms are like, you know, literally a meter squared or whatever. I get in, close the door and the toilets, like, literally whacking the knees. And it's the bits that I take for granted at home. Grab bars, you know, the toilet, which you know. Something I don’t want to talk too much about, but it's, you know. And so it's it's not that I'm trying to age myself and not go out and do things or or restrict myself, should I say. But the creature comforts at home”.<br><br>PwP (T6 Complete, Male): “Any payments. We’d be better going the road than on the pavements because it's not. Yeah. And sometimes they just drop like that and you hit them and then that's it”.<br><br>PwP (L1 Incomplete, Male): “And when you find anything like this on the road, you end up in the in the road because the pavement slopes like that *laughs*”.<br><br>PwP (T12 Incomplete, Male): “Yeah, it's solving my problem with the van. Tail lift, I'm not on a path anymore, yeah. I'm coming onto the road cause the lift comes out the back of it, right. And then I'll have to try and get up curbs. I've just had to pay £1400 to get a drop curb, which I have to pay for in the road. And you have to apply for it as well. Disgusting. And everyone else can use it. But I have to pay for it”. |

| | | | | |
|---|---|---|---|---|
| | | | | PwP (T6 Complete, Male): “We weren't allowed disabled parking”.<br><br>PwP (T4 Complete, Male): “Yeah, some limitations like that, as Leon said. You go to someone else's house. Different ball game, right? Less manoeuvrability, more furniture. Layout”.<br><br>PwP (L1 incomplete, male): “Non-accessible space, I guess. Or non-adapted space [is a barrier]”.<br><br>PwP (T12 Incomplete, Male): “Going shopping, right? Can't get this one. You have to look around for a member staff, the hardly like Tesco's, there's hardly any staff and you gotta wait for someone to come down the aisle “Excuse me, I don't suppose you can get that”. I hate that. I like to do everything as much as I possibly can, but with certain jobs you can’t. You physically can’t do it”.<br><br>PwP (T9 Complete, Female): “It’s worth mentioning, you know, depends where you go, if you need to use a lift and the lift isn’t working”.<br><br>PwP (L1 Incomplete, Male): “Yeah, right. And if that's something that gets you out of your house, you can't then do anything else because you're lift is broken”.<br><br>PwP (T12 Incomplete, Male): “My sports centre as well. I joined the gym there and the lift was always broken. Got there - broken. Have to go home without it. Disappointment”.<br><br>PwP (T4 Complete, Male): “Again, it's postcode isn’t it. Where I've moved to, there’s a hotel next door to where we live, now. They've got a leisure facility. I rang them up and said “is the equipment in your gym wheelchair accessible?” and they said “of course it is, we've had a we've had a a multimillion |

| | | | | |
|---|---|---|---|---|
| | | | | refurbishment last year". I went "brilliant". So I go down there, make an appointment to see the membership manager, he takes me into the gym, and none of the equipment is accessible. So OK. All the machines are all fixed and he went "Oh, I know it's all fixed" and I said "well I wouldn't even bothered coming here if I was told it was not wheelchair accessible". And the very fact that a good hotel has spent that much money recently, and not made it accessible equipment, is just".<br><br>PwP (T12 Incomplete, Male): "I'm out eating, and needed a pee. I go "where's the toilet?", he says "upstairs". I go "where's the lift?", he says "we ain't got one? They put a disabled toilet upstairs just to say they've got a disabled toilet. You can't use it in a wheelchair".<br><br>PwP (T4 Complete, Male): "Just going back on to sort of like diet and stuff like that, a lot of people refer and you sort of like the Apple Watch. "We've burned this amount of calories" and stuff like that, but that's not tailored to someone just using their arms, if that makes sense. Yeah, there's nothing that sort of like tailored to a paraplegic. Like, if I cycle on my bike, yeah, my heart's running at 140 beats a minute or whatever for 2 1/2 hours, it will stay all I've burnt X amount of calories, but that's based upon the calculations that it would be for my quads and my legs drawing that energy and not my arms. So, people think, "oh, I've burnt all this energy", whereas in fact you haven't because what you're using is nowhere near the burn. Does that make sense?".<br><br>HCP (Physiotherapist, Female): "If we also look at the capability of some of our patients being able to break some of that very static behaviour and we're not necessarily looking at exercise or intense activity, the home environment is also really key factor because a lot of our patients will sometimes get discharged to a |

| | | | | |
|---|---|---|---|---|
| | | | | microenvironment. So even just being able to manoeuvre their wheelchair around will be sometimes extraordinarily difficult to do so breaking up any activity may be extraordinarily difficult because they may not be able to manoeuvre even from room to room”.<br><br>HCP (Physiotherapist, Female): “Lots of people are discharged to live in one room [with little space to move around and do activity]”.<br><br>CCG (Charity worker, male): “When they're discharged from hospital now aren't always going home because their home environments aren't accessible… So it's certainly stuff like that is going to affect their ability to be active”.<br><br>CCG (Family member, Female): “So for example the OTs couldn't get a standing frame for over 12 months. She's only just got a standing flat frame for her to be able to use. It's taken a long time for the external community occupational health to get involved. And just mimicking what the other guys have said in regards to the home environment she was popped into a living room area with a minimal access to upstairs, in fact had no access to upstairs for obvious reasons, but she had minimal access to her bathroom. It was a case of discharge and hope for the best effectively. So she had to get a kind of a mini. Well, she had to convert part of the house, which isn't ideal, but into almost a wet room type facility for her to be able to get herself washed and dressed”.<br><br>PwP (T12 Incomplete, Male): “Same as opening the fridge, you're doing so many bloody moves. Just to open the fridge door”.<br><br>PwP (L1 Incomplete, Male): “Yeah you have to get clearance or. Yeah, the dishwasher and like our units in the kitchen are |

| | | | | |
|---|---|---|---|---|
| | | | | kind of like this. And the dishwasher is opening here. I can I can get some stuff this way, but if I start a job on the other side, I'm kind of stuck on that other side because the door's open. So I can't get this stuff out to put in the cupboards because they're backside sort of thing”.<br><br>PwP (L1 Incomplete, Male): “Yeah, space. Physical space [is a barrier]”.<br><br>PwP (T12 Incomplete, Male): “And if you’ve got stuff at home, you can do it yourself. But a gym in your house, you might not have enough space. You’re doing it on your own, you don’t. You hang stuff on it. You don't even use it. It's just sat there doing nothing”. |
| | | | Cost and lack of access to equipment to do physical activity or exercise (barrier) (n = 10) | HCP (Physiotherapist, Female): “They do normally go home with an exercise plan and we can send them home with the Theraband. We wouldn't send them home with weights, they'd have to self-purchase. And the thing I suppose, all the all the the bikes or additions to their chair, they would have to self-purchase”.<br><br>CCG (Charity worker, male): “There's not a lot that you can get that is complimentary. You know, a lot of this stuff now you've got to pay for, and it's not cheap”.<br><br>CCG (Family member, Female): “So for example the OTs couldn't get a standing frame for over 12 months. She's only just got a standing flat frame for her to be able to use. It's taken a long time for the external community occupational health to get involved. And just mimicking what the other guys have said in regards to the home environment she was popped into a living room area with a minimal access to upstairs, in fact had no access to upstairs for obvious reasons, but she had minimal access to her bathroom. It was a case of discharge and hope for the best effectively. So she had to get a kind of a mini. Well, |

| | | | | |
|---|---|---|---|---|
| | | | | she had to convert part of the house, which isn't ideal, but into almost a wet room type facility for her to be able to get herself washed and dressed".<br><br>PwP (T4 Complete, Male): "It's getting dressed. But I was also thinking more just to the exercise. You know the getting big physical, doing physical activities, being non-sedentary which, is a lot, again it's easier because it's all there. Yeah, it's like 2 minutes away rather than "oh I've gotta get to the gym" or "I've got to go for a walk" or "I've got to do something in the park" or whatever".<br><br>PwP (L1 Incomplete, Male): "It's all so defeating, isn't it? Really? I think that's what I'm saying. The hardest bit for me is getting in the car to go to the gym".<br><br>PwP (T12 Incomplete, Male): "That's the only bit of kit I got from the hospital. A rubbish wheelchair and a standing frame".<br><br>PwP (T4 Complete, Male): "I want a home bike again".<br><br>PwP (T4 Complete, Male): "Yeah, exactly. So so some stuff you know. And then I had approval for NHS funding for my standing frame. But my case manager said it's probably easier to wrap it all up in one funding claim against the, you know, the insurer. So it's all, it's complicated".<br><br>PwP (T6 Complete, Male): "Or your equipment that you can get to [affects opportunity to break up sedentary behaviour with physical activity]".<br><br>PwP (T4 Complete, Male): "So instead of steps, it measures pushes. "Yeah. Brilliant". Yeah, and it does, but it still doesn't track your distance. So if you go into your health app and it will say "you've done 10,000 pushes today". And you go "Well, how |

| | | | | |
|---|---|---|---|---|
| | | | | far have I gone?”. It will say “you’ve gone 0” because you haven't taken a single step”.<br><br>PwP (L1 Incomplete, Female): “Yeah, there's no accessible gym where I live”.<br><br>PwP (T11 Incomplete, Female): “The availability. Yeah, just the availability to services really or like the the access, you know somebody might not live in an area that has an accessible gym, maybe not have access to the Internet. I mean that would be odd these days. But you know, I think just what people's, you know, availability to services or places would be”.<br><br>PwP (T4 Complete, Male): “Yeah, I think if you look at them they’re all sort of like in inter-linked with each other. Yeah. And like availability, and like equipment at home that you spoke about. I'm very fortunate that I've I've retired from work and I can spend time on my body, which is my job. My job is my body and looking after myself. I've got, I've got an FES machine upstairs, which does my legs. So I put like for two hours every couple of days, I FES my legs. Yeah, to keep bulk, keep muscle, keep the pain out, keep the lactic acid out, but I’m fortunate that. That bit of kit is like 12 grand. Yeah, I’m fortunate that I've got that at home to use. Yeah. And I would do that for two hours every couple of days. Something like that should be given to every paraplegic person who it’s suitable for, but it's not, and that's what’s not right”.<br><br>PwP (T4 Complete, Male): “You might you might get a set of ramps on your own from your OT if you’re lucky”.<br><br>HCP (Physiotherapist, female): “They haven't got any finance finances available to purchase any extra equipment and that might be what's needed to be active”. |

| | | | | |
|---|---|---|---|---|
| | | | | PwP (L1 Incomplete, Male): “Yeah, exactly. Sometimes my wife gives me a once over. She.. in terms of looking for for things. It's less, you know, she’s less attentive now 14 years after the accident. But the the talking about physical exercise or what have you. When I was discharged from hospital and I had my insurance covered like a physical person, trainer, whatever coming in a couple of days a week, and then it got to a point, basically the insurance was saying “you're not making anymore progress”. Yeah, you are maintaining your level of exercise. Yeah. “So, we can't pay for this anymore”.<br><br>PwP (L1 Incomplete, Male): “So we paid for me to go and see someone a couple of days a week out of our, you know, pocket, which was great because I didn't wanna stop doing the physio or being in a good sort of routine and doing as much as I could”.<br><br>PwP (T6 Complete, Male): “It's not titanium [wheelchair], so I can do what you were saying. Yeah, take the wheels off when you get to the car, put the wheels in the back and then this goes in the passenger seat. So I can go out independently. Umm, but the more you have taken off the chair, the more the price goes north”.<br><br>PwP (T6 Complete, Male): “14 years. Yeah, but you then have to bite the bullet if you can. If not, because luckily enough, I was lucky to get through government funding and work funding. Otherwise I'd still be in one of those [heavy wheelchairs]”.<br><br>PwP (T4 Complete, Male): “I will have to pay for it [equipment needed to be active] myself”.<br><br>PwP (T6 Complete, Male): “So, you know, it's what PwP is saying, if you don't have money, you are absolutely stuffed outside [of inpatient rehabilitation]”. |

| | | | | |
|---|---|---|---|---|
| | | | | PwP (T4 Complete, Male): "It's just a you have to navigate the right way. You have to get the right budget approved, some of its NHS funded. Some of it has to be privately funded".<br><br>PwP (T4 Complete, Male): "And one of the biggest barriers is money. Yeah. I'm fortunate that I can afford to buy a new wheelchair. I can afford to buy it and buy that and have a very active life. But some people can't. Yeah. And that's where it goes wrong. Is that people who are walkers, yeah, don't seem to understand… And that's what's that's what's wrong, about picking up what Donna said, is that you don't understand. Not you. Anyone, unless you're in this position and it's wrong that finance should be a barrier. And but it is a barrier for people. And that's not fair".<br><br>PwP (T4 Complete, Male): "It's in the ability that if you haven't got to work because you you can financially rely on your pension or retirement, yeah. Then, instead of working 9:00 till 5:00, my body's my job from 9:00 till 5:00. So when my son went to school, yeah, I'm lucky that I didn't have to go to work. So he went to school. I went to work on my body. So when he come in from school, I could do anything with him because I was physically fit enough to do that, if that makes sense. So that's a barrier for people as well".<br><br>PwP (T4 Complete, Male): "Well, yeah, again that comes down back down to the old finance issues, doesn't it? Yeah. Are you able to buy a house or put an extension on or adapt your house to support the needs of a wheelchair? And that unfortunately costs money, which is wrong because you won't get nothing from the government". |
| | | | Wheelchair being inappropriate for physical activity (barrier) (n = 7) | HCP (Physiotherapist, female): "They get given much heavier chairs. Harder to propel and definitely harder to do wheelchair skills in". |

| | | | | |
|---|---|---|---|---|
| | | | | HCP (Physiotherapist, Female): “So there isn't a circumstance in which somebody goes home unable to mobilize. That's unthinkable to us. That's not true in all the spinal cord injury centres and in an increasingly, because of the difference between what the wheelchair services provide and what our patients need. The reality is that gets closer with every passing month. But, as HCP said before, it's not necessarily you know they'll learn to function at a really high level in an appropriate chair for their needs here. And they will almost always be discharged in something that is vastly inferior, which will mean pushing is heavier, more painful, they're sat less well, they get less, they're less comfortable in sitting, they can access the environment less well that you know, the transfers are more difficult. All of these things, because the first chair that they get from their wheelchair service is rubbish”.<br><br>PwP (T4 Complete, Male): “Yeah, I think there's there's an awful lot of problem solving that we… from dishwasher, cupboards, you know, whether it's going to the toilet camber on the pavement. Yeah. So I'm always thinking about what is that thing in front of me and how do I kind of best position myself to go around. And now I'm I'm waiting for a better wheelchair, for example. So it's really funny the NHS, brilliant. But you know”.<br><br>PwP (L1 Incomplete, Male): “I'm having problems. This will chairs not been looked at, serviced, since before covid and it’s falling apart”.<br><br>PwP (T4 Complete, Male): “You get a super lightweight, active user wheelchair at the spinal unit, which you practice in and practice in and practice in, and then you get a going home wheelchair”.<br><br>PwP (T12 Incomplete, Male): “You can’t get it out of the car. When you pass your car test, yeah, you're supposed to |

| | | | | |
|---|---|---|---|---|
| | | | | dismantle your chair and pull it over you, right. I knackered my shoulder doing that. Now I've got van now, with a tail lift, yeah. I was supposed to drive from a wheelchair, but it's only if it's a electric wheelchair, which I can't light as well. So I said “I'll have an electric wheelchair, then”. So I was gonna buy an electric wheelchair just so I could drive my van”.<br><br>PwP (T4 Complete, Male): “At the moment kind of just moving around the house is more difficult in this wheelchair”.<br><br>PwP (T6 Complete, Male): “I spent ages in one of those [wheelchairs] and it nearly killed me”.<br><br>PwP (L1 Incomplete, Male): “My wheelchair was looked at in February because they're still waiting for cushions, parts. Yeah, it's all the duct tape, the little things inside are like pyramid-shaped with air. But last time I looked, because I've washed them all, most of them are flat. Uh, I took the breaks off as they were hitting my hands”.<br><br>PwP (T4 Complete, Male): “And this, just to pick up what PwP just said there about picking the chair up. This comes back to that we've got to fight for everything. Yeah. So, for instance, my wheelchair. I'm very active. I'm in my own wheelchair that I've bought. Wheelchair services, yeah, wanted to give me an Argon 2 wheelchair that weighs a bloody tonne, yeah, and lifting that in and out of my car with my shoulder is no good whatsoever. Yeah. So it's just pointless. I can't use it”. |
| | | | Geographical inequalities to services or opportunities relating to physical activity and exercise (barrier) (n = 5) | PwP (L1 Incomplete, Male): “I think financial, but also geographic. Yeah, I think probably, yeah. Postcode kind of lottery. Yeah, you might get better care in certain places”.<br><br>PwP (T6 Complete, Male): “It depends on the hospital you end up under as well. The hospital you end up under, because I wasn't under that spinal unit [that was previously mentioned]. |

| | | | | |
|---|---|---|---|---|
| | | | | And then listening to what you lot have been given coming out that spinal unit. And I was in another spinal unit".<br><br>PwP (T6 Complete, Male): "With a sub group because as we said postcode lottery was that it. If you get too big and too, then you're like, some people will be like "well, why they getting that? We're not getting that" and vice versa".<br><br>PwP (T9 Complete, Female): "Perhaps distance. Because you know, because I want to take up fencing. But it's far too far away".<br><br>PwP (L1 Incomplete, Male): "Yeah, yeah. Location of the activities [is a barrier], you know".<br><br>PwP (L1 Incomplete, Female): "Yeah, there's no accessible gym where I live".<br><br>PwP (T11 incomplete, female): "The availability. Yeah, just the availability to services really or like the access, you know. Somebody might not live in an area that has an accessible gym". |
| | | | Support from the workplace (facilitator) (n = 4) | PwP (T6 Complete, Male): "I was fortunate enough to get obviously a donation through whatever they are, the government thing and then work pay for this chair".<br><br>PwP (T12 Incomplete, Male): "Actually they do, don't they? If you work, they pay for loads of stuff."<br><br>PwP (T11 Incomplete, Female): "I think it's the type of job that you do really. I mean, I'm a nurse at a nursing home. That's my job, so I'm constantly moving".<br><br>PwP (T11 Incomplete, Female): "Not really. I mean, I'm. I'm going across the nursing home. I'm either in meetings, you |

| | | | | |
|---|---|---|---|---|
| | | | | know, meetings, but I'll be wheeling myself to and from external buildings. Yeah. Transferring in and out. Transfers onto the toilet and back off again".<br><br>PwP (T4 Complete, Male): "I adapted my workspace and my work were very good with me in providing that sort of stuff".<br><br>PwP (T4 Complete, Male): "100% we have. So you got you get an assessment, an independent assessment and you say what you need and they [your workplace] have to supply these things". |
| | | | Excessive planning and inability to be spontaneous (barrier) (n = 3) | HCP (Physiotherapist, Female): "Or the uncertainty. So, everything has to be planned very differently from most most people who have different levels of mobility. So you can't be, you have to be very brave and very able to be spontaneous. And so if you're with a group who where normally it would be completely spontaneous where you ended up, there are some people who are chair users or who are ambulant post their cord injury, who are really comfortable with that, but it's a tiny minority. Because you have to, you know, you plan where you're going, you know if you can access it, you know if you if there's enough room".<br><br>PwP (T6 Complete, Male): "The amount of planning you have to do just to go out the door is sometimes unbelievable".<br><br>PwP (T9 Complete, Female): "You've got to go through the rigmarole of, you know, what assistance do you need [when going out and about]".<br><br>PwP (T9 Complete, Female): "Oh well I got on a tube to come off at this stube station, and they asked "do you need a ramp to come off at Uxbridge?", so "yes", somebody phoned ahead. [I arrived and] it wasn't there". |

| | | | | |
|---|---|---|---|---|
| | Social opportunity | Social influences | Support from family or friends (facilitator) (n = 11) | HCP (Physiotherapist, Female): “Yeah, we occasionally see some families that are very much facilitators though as well in terms of let’s do your exercises, let’s do your standing frame and they do, it's it's probably less, but they do push potentially, so they they could be classed as a facilitator as well as a barrier”.<br><br>HCP (Physiotherapist, Female): “In my experience, my patients I see as outpatients that have got that support network definitely go out and about an awful lot more than the ones that potentially don’t. Or have a close family unit that will support them. It does make a big difference in in my experience”.<br><br>CCG (Charity worker, Male): “Well, if you're a parent with young kids, it's gonna be. It might be, I suppose you're not likely to be that sedentary in some in some respects, but but again, if you've got, you know, home commitments and things that you need to be doing in that respect. Then it can be a a distraction, I suppose from focusing on on being active in in some ways. Yeah, but there's. It depends, I suppose, whether or not. Yeah. You know, you might be a mother with young kids or, yeah, the people have different family commitments, don’t they, which can be a distraction from from engaging in some kind of exercise or activity”.<br><br>CCG (Family member, Female): “I think the kids, my sister [with paraplegia] has got two, in fact she’s got four children, but two younger children and she's she, it keeps her quite busy”.<br><br>CCG (Family member, Female): “I think you know I I try and encourage her to go out for some we call it wheels but walks effectively. So we'll go out for a walk and a wheel together”.<br><br>PwP (L1 Incomplete, Male): “Having two daughters and a wife, sedentary behaviour is just not allowed, which is good, which is |

| | | | | |
|---|---|---|---|---|
| | | | | good. They never allow me to wallow myself in self-pity or lay on the sofa”.<br><br>PwP (T12 Incomplete, Male): “I get on the bed at night. Yeah, and just watch film after film after film after film. Get up, have a bit of supper. Go to bed, go to sleep. Wake up in the morning. Busy, busy, busy, round my mum’s sorting her house out, cos my daughter's gonna get it eventually. So we're trying to decorate it, plasterboard it, and do everything. which I take part in. I do all that sanding and stuff like that. But then at night when I get home, after my tea I get on the bed and couch”.<br><br>PwP (T4 Complete, Male): “I think that's similar to what I do. In my workshop, I restore things, up cycle things. Yeah. And so and also my wife gets me doing the DIY jobs which, it's amazing what she’ll let me do when it's a DIY thing”.<br><br>PwP (T12 Incomplete, Male): “No, I love staying in, if that mean’s I have to stay in. On a Sunday, if I don't go to my mum's, that's it. My day is literally on bed all day”.<br><br>PwP (L1 Incomplete, Male): “Yeah, because they're cause. You're you're. Yeah, you're stopping them [children] from rolling away that way. Or toddling”.<br><br>PwP (L1 Incomplete, Male): “Right, but it, but if it's getting up a ladder or going up into the loft. Yeah, to help my wife organize it, she's alright with that. My wife is happy for me to climb up a step ladder to get into the loft to help her organise the loft. It's cause she wants that job done”.<br><br>PwP (L1 Incomplete, Male): “Yeah. And I and I guess that's that's what you sign up to as a parent. So you know. They they do sort of distract me from whatever I'm doing for whatever they |

| | | | | |
|---|---|---|---|---|
| | | | | need to get done, and I'm not complaining about it because I love my daughter, I love my wife”.<br><br>PwP (T6 Complete, Male): “Yeah, I guess family depending on who they were, going back to my last lot, whether it's parents or whatever. It's dependent and you go from chalk to cheese. My son would just say “get on with it”. And others would not let you get anywhere near it”.<br><br>PwP (T4 complete, Male): “There's a thing, isn't there? About community, right? So doing things in a solitary capacity is often quite hard. A lot of barriers to it”.<br><br>PwP (T4 Complete, Male): “Yeah, you know, then again, that community aspect is so encouraging, finding ways for people to connect, cause that”.<br><br>PwP (T1 Incomplete, Male): “My sons are as like I used to be as a teenager, they're 15 and you're going on the bench press going “I did 50 kilos yesterday” and there's twins, so the other one is going “well, I benched 55”. So just yesterday they both bench pressed 55 and and that's wonderful. But you see what I mean? You got to start. I said, why don't you do reps and then build your way up and, you know, don't go for the powerlifting 100 kilos. You can see what I mean? So you know it's it depends on the person's incentive”.<br><br>PwP (L1 Incomplete, Female): “Well, I don't do the cleaning. I have someone comes in, does the cleaning for me. And while they're there, they supervise me, doing like a batch cook for the week, and then I just reheat in the microwave every day for what I'm eating once a day. And then I do the washing when they're there as well, 'cause. Then they can get the stuff in and out the washing machine for me”. |

| | | | | |
|---|---|---|---|---|
| | | | | PwP (T11 Incomplete, Female): “I don't sit still long enough. I also have five children so I can't sit still for very long periods of time”.<br><br>PwP (T1 Incomplete, Male): “Yeah, actually, friends and family, it'd be a good idea to help build to motivate them”.<br><br>PwP (L1 Incomplete, Female): “Yes, because [having someone to do activity with] it would encourage me to go out more”.<br><br>PwP (T4 Complete, Male): “I think that's very unique to the specific person and what that person needs are me personally, I wouldn't need that. Some people might need that and therefore I can be a friend, a sponsor or something like that, and they can help together to do it until that person can do it on their own, if that makes sense. So have someone to support you to do it until you can do it on your own”. |
| | | | Being deterred by family or friends (barrier) (n = 8) | HCP (Physiotherapist, Female): “I think it depends on like where they’re discharged to as well and how much support they have at home, because like even those insignificant movements, a lot of patients won't do them because they have other people there to do them for them. And I think like a natural human response to someone who's less able than you are, is to help them. And so family and other people around would tend to just do small things like going out to get a cup of tea instead of them moving into another room to do it themselves. Things like that. So it depends on if someone is living on their or if they’re living with four other people that can help”.<br><br>CCG (Charity worker, Male): “Well, if you're a parent with young kids, it's gonna be. It might be, I suppose you're not likely to be that sedentary in some in some respects, but but again, if you've got, you know, home commitments and things that you need to be doing in that respect. Then it can be a a distraction, I suppose from focusing on on being active in in some ways. |

| | | | | |
|---|---|---|---|---|
| | | | | Yeah, but there's. It depends, I suppose, whether or not. Yeah. You know, you might be a mother with young kids or, yeah, the people have different family commitments, don't they, which can be a distraction from from engaging in some kind of exercise or activity".<br><br>CCG (Family member, female): "I find my mum does a lot for him and maybe that stops him from doing things himself".<br><br>PwP (T4 Complete, Male): "I think that's similar to what I do. In my workshop, I restore things, up cycle things. Yeah. And so and also my wife gets to be doing the DIY jobs which, it's amazing what she'll let me do when it's a DIY thing, but if I wanted to do something, then "oh no, no, no, that's far too dangerous. No, you can't be doing that"".<br><br>PwP (L1 Incomplete, Male): "Yeah, definitely. Because I gave the example. And like is, is this sort of hypocrisy that my wife shows that like, if I said "well, I wanna go watch Crystal Palace play" she would say "well, how you going to get there? Is there parking?"".<br><br>PwP (T12 Incomplete, Male): "Yeah, they [family] put barriers up".<br><br>PwP (L1 Incomplete, Male): "No, and I I have always appreciated that. There's not, because that's the flip side to it. You go to people's houses or other relatives that don't see your day to day, so they don't really know what you're capable of and not capable of. And they sort of try and wrap you in cotton wool".<br><br>PwP (L1 Incomplete, Male): "You know, it's like:<br>"Ohh can you do that? Can I get that for you?". Sometimes I quite like it, like please do grab me that and do that. Yeah, |

| | | | | |
|---|---|---|---|---|
| | | | | yeah, yeah. I'm like "this is fantastic", but equally it can annoy you".<br><br>PwP (T12 incomplete, Male): "Family can put barriers up for you. But if you got your own barriers, so it's two fences to get over".<br><br>PwP (L1 Incomplete, Male): "You want them to be going "This is a fantastic opportunity. Can you imagine how you'd feel if you've got this done?". You'd have been six weeks since South Africa, You'd have survived all shootings and everyone trying to kill you, but you'd have your pilots thing. And for anyone that's a massive achievement, you know, and you sort of think "you should be behind me, guys, not sort of pointing out all the potential dangers, dangers""<br><br>PwP (T4 Complete, Male): "So for example, and again it's a relatively new and I new for my wife as well. I don't forget, but yeah, we just moved to a new area. New-ish as in sort of quite quite close to where we used to live, but it's still a new town. And she was like "oh well before we go to that pub, let me go first and check it out. Make sure it's all OK". And yet she was quite happy for me to come to Brunel University on my own, you know, and just getting a cab and come here and find my way to the right building and find my way in. And then yet the pub that's two doors down to where we live, whereas I was saying to her "Well, let's just go and yeah"".<br><br>PwP (T6 Complete, Male): "Yeah, I guess family depending on who they were, going back to my last lot, whether it's parents or whatever. It's dependent and you go from chalk to cheese. My son would just say "get on with it". And others would not let you get anywhere near it". |

| | | | | |
|---|---|---|---|---|
| | | | | PwP (T4 Complete, Male): "There's a thing, isn't there? About community, right? So doing things in a solitary capacity is often quite hard. A lot of barriers to it.<br><br>PwP (L1 Incomplete, Female): "Umm. Well, I don't have much contact with my family. I mean, I go for dinner with my parents once a week. And that's it. And I've only got 2 friends and I see them once a month, so I don't have that support there to give me the incentive". |
| | | | Peer support network (facilitator) (n = 7) | HCP (Physiotherapist, Female): "But also this might be quite interesting in terms of the difference between a spinal unit and being treated somewhere else in a non-specialist centre, because of a lot of our patients will develop relationships with people that are on the ward themselves. And again, not always, and I'm generalizing a little bit. But then the patients will go out and do things together. So again, they've got those peer support networks to ultimately, again, together overcome some of these barriers".<br><br>HCP (Physiotherapist, Female): "And they'll exchange numbers et cetera. So, so it depends who's on the ward at the time as well. Some of them develop really close friendship groups that last a very long time".<br><br>HCP (Physiotherapist, Female): "In my experience, my patients I see as outpatients that have got that support network definitely go out and about an awful lot more than the ones that potentially don't. Or have a close family unit that will support them.<br>It does make a big difference in in my experience".<br><br>CCG (Family member, Female): "I think for my dad, he doesn't really, you know, sport has never been part of his life. He doesn't really have contact with other people with a similar injury. I think he just feels quite isolated. I think to have |

| | | | | |
|---|---|---|---|---|
| | | | | something like that where he could see other people actually are able to have some level of activity. I think really benefit him".<br><br>CCG (Charity worker, Male): "If in some way it can involve contact with others, I think that that's one of the best motivational things out there".<br><br>CCG (Family member, Female): "I was just thinking, you know, I could be added benefits as in sort of social, psychological, I think definitely for my dad. Because of his age, he does feel quite isolated with with his injury. And yeah, I just it could have. Yeah, more than just physical benefit for him. I think it could. Yeah, actually, something like that could actually really improve his life in lots of ways".<br><br>PwP (T4 Complete, Male): "There's a thing, isn't there? About community, right? So doing things in a solitary capacity is often quite hard. A lot of barriers to it. You know, whereas, you know, if you're doing it with a buddy, or better still, with like, you talk about your walking could group or standing group".<br><br>PwP (T4 complete, Male): "Yeah, you know, then again, that community aspect is so encouraging, finding ways for people to connect".<br><br>PwP (T6 Complete, Male): "Along those lines. At rugby, 15 of us, however, many of us, 16. And someone said like quite a while ago, "there's sixteen of us here and we're all talking to each other now. If it wasn't the fact that we were sitting in this chair, we wouldn't be talking to each other". We would probably have nothing to do with each other. Walk past each other on the street because our interests probably lay in different ways. But the fact that we're in this chair brings us together and gets us going, doing different things. Yeah, so that kind of makes it". |

| | | | | |
|---|---|---|---|---|
| | | | | PwP (L1 Incomplete, Male): “And it's funny because isn't it? It's still the people say that in the negative way, so it's like just because I'm in a wheelchair doesn't mean I'll get on with like the person who's in the wheelchair. But having that common bond and under. It's an understanding which you don't need to describe to another person in the chair because they know exactly what you're talking about. Yeah, the the different things, it's just sort of a telepathic understanding”.<br><br>PwP (T6 Complete, Male): “the pros and cons, and whatever you wanna call it. Yeah, the natural side of it just comes out and I mean, if you look at the five of us sitting here now, never met each other, but yet we’re properly just bleating our hearts out. Get what I mean?”.<br><br>PwP (T4 Complete, Male): “It’s what. It's not. So this is, socialization, right? Which is usually helpful, but I think it's more that like the spinal charities try to organise this. It’s kind of community. It's about. But I've only been in my new town for two- and a-bit weeks. I've not seen a single other wheelchair user”.<br><br>PwP (T4 Complete, Male): “I would like a way to be able to connect with other, you know, wheelchair, or, even better, spinal patients”.<br><br>PwP (L1 Incomplete, Male): “So if all of these users are on this app, they're all spinal cord injury patients. They're all trying to be more active and less sedentary, and I don't know if, it's not like goal setting, or it's like “Mark's smashed this target” or “he's achieved this this week” and “so and so on has achieved””.<br><br>PwP (T6 Complete, Male): “Or there's a. I mean, for, depending on what state you’re at. So for somebody like PwP, obviously everything's very new. So for something like, if you have a |

| | | | | |
|---|---|---|---|---|
| | | | | group, if need be, and if he, is obviously different ones wants to do it in different ways, but he would learn so much for. Obviously everyone that’s been there for a year, six months or 15 years, because everyone's at different stages and everyone tackles things in different ways. So what works right for PwP, and what works right for PwP, and what works right for me… [might be different].<br><br>PwP (T4 Complete, Male): “But you did say that meeting your wheelchair rugby buddies is hugely helpful”.<br><br>PwP (T4 Complete, Male): “No, I was thinking more of it, in my local communities like you know. Tt's like having a running buddy or a gym buddy. You know, it's just somebody you go “You know what? Let's go. Let's go and do that together””.<br><br>PwP (T6 Complete, Male): “I was more curious as to going back to the rugby, I've never done anything like that before. It was more curiosity to me that got me doing that type of thing. And then it led into the.. we go training once a week. So it's the physical side of it, but. Yeah, and then the community side of it and there's. And then you might stay behind after and just have a cup of tea or something and just a yarn. It's just silly little things, and then”.<br><br>PwP (T6 Complete, Male): “The enjoyment bit umm, I was thinking maybe that would be linked to community, in terms of if you're feeling a little bit demotivated, you know, maybe there's like a chat or a forum or something. Maybe. You could go “I’m feeling a bit low about this or that”. I don't know. It's not something I would probably do”.<br><br>PwP (L1 Incomplete, Female): “Yeah, that would be good. I use WhatsApp. If there was a WhatsApp group [with other individuals with a spinal cord injury] that would work”. |

| | | | | |
|---|---|---|---|---|
| Motivation | Reflective motivation | Beliefs about capabilities | Low self-esteem or self-conscious (barrier) (n = 8) | HCP (Physiotherapist, Female): “Yeah, just a couple of other bits on that. Another thing is confident, so feeling confident to get out and about. And again, if you for us, we're talking about a population from people who are babies to people who are first injured in the 80s, so their needs from an activity point of view are different um and, but their ability to get out of the front door and access outside space and and be happy moving around is really variable. So confidence to access business”.<br><br>HCP (Physiotherapist, female): “People don't like being seen in a wheelchair”.<br><br>CCG (Charity worker, male): “Psychological components around motivation and confidence and self-esteem, body image. That that can all contribute to just not finding that 5 minutes [to be active per day]”.<br><br>CCG (Charity worker, Male): “Yeah, I I think that. I think that's a huge thing. For a lot of people a big issue for them is is being seen as being disabled. You know, being. The chairs are a very visible cue, obviously, and some people find it, yeah, really, really hard to adjust to that”.<br><br>PwP (T12 Incomplete, Male): “People staring at you sometimes puts you off as well doing anything”.<br><br>PwP (T12 Incomplete, Male): “People talk about you, nudging, winking, everything, pointing [when going out and about]”.<br><br>PwP (T6 Complete, Male): “Or it’s easier because I’m in the chair and when they come out with me and I've got the dog and the chair, they say “I don't know how you managed to get round the park”. Because everyone wants to stop and talk to you, or help you, or wipe your backside for you, so to speak, because you're in the chair. It's just, I don’t know. Leave me alone”. |

| | | | | |
|---|---|---|---|---|
| | | | | PwP (T6 Complete, Male): “You feel like a three-year-old going shopping because you can't reach above that level”.<br><br>PwP (L1 Incomplete, Male): “Yeah definitely, like lack of self-esteem or something that could come from the injury, illness”.<br><br>PwP (T6 Complete, Male): “Yeah if you think you can't do it, you don't. Some people won't even try it”.<br><br>PwP (L1 Incomplete, Male): “And yeah, with that, it could be other people's opinions on what you're doing and criticism. People might be criticising what they see is a pathetic attempt to to do whatever”. |
| | | Intentions | Building habits around activities of daily living (facilitator) (n = 7) | HCP (Physiotherapist, Female): “Most paraplegics will be physically able to do that [do activities of daily living]. In fact, I mean almost all paraplegics, well there might be if they had something else wrong with them, then maybe that would mean that that was technically not possible, but with a paraplegic level of injury in a reasonable environment you should be able to independently get out and about. It's the environment that prevents you doing it, not your intrinsic skills.<br><br>PwP (T11 Incomplete, Female): “Yeah. So I am, you know, with a spinal cord injury, you have. I have to use catheters to go to empty my bladder. So I will deliberately get up, go over to the toilet rather than using bags. That's something that we were that. That’s a way for me to be able to kind of move or make me make myself a cup of tea. I might get up out of the chair to do that, but I do a lot of transfers so I don't stay in my chair very long. I will transfer out of my wheelchair onto the sofa or, and out to the kitchen. And so I think I just. I don't sit still long enough. I also have five children so I can't sit still for very long periods of time”. |

| | | | | |
|---|---|---|---|---|
| | | | | PwP (T1 Incomplete, Male): “And you can't be embarrassed about your own incompetence. I'll be on that after this. But that's the truth of it. So yeah, building it in and building it up at home, piece by piece, that's a good idea. You're like lifting tins or lifting coffees and things, and that simple stuff. Sitting on the sofa squeezing those”.<br><br>PwP (T1 incomplete, Male): “Building [activity] into things you've got to do in the house, it gives you a sense of worth, not just for exercise and fitness”.<br><br>PwP (T1 Incomplete, Male): “But the. But lifting the tin of beans there, just doing what you can, you know, get hoovering or something. That's a good idea [to build activity into daily life]”.<br><br>PwP (T1 Incomplete, Male): “And so there's, you know, so there's the things you gotta get the I think the intellectual problems, you know that the the thought process then the physical aspects and then slowly but surely you you knock every little brick down and work on it. And you know, then there's it's just, I think. I really like your idea about the exercising at home. Just build it into your lifestyle because unless you're just going to let, and even if you're laying in bed, I have got grab rails at the side because I have no stability, and I I actually have very strong arm, but I I grab them and I pull myself across the bed. And then I use gravity to put me in the wheelchair. I've had my bed raised above the wheelchair to just fall into it. It's a little more fun getting back in again. But if that happens, I just end up lying on the sofa all night. You know that's it's the way. But you see what I mean. There's always ways of building some form of exercise into your, into everything”.<br><br>CCG (Charity worker, Male): “If I if I'm watching TV now, I will. I won't sit there for, I'll make sure I'm not sat sedentary for more than an hour. For example. You know, if I've seen an hour come |

| | | | | |
|---|---|---|---|---|
| | | | | up, I'll make sure I get up. Even if it's just to go to the toilet or go and just do something else and then sit there. It means having to transfer back into my chair and to move. But but. But things like that. I think in some ways getting a bit of a trying to get a bit of a timetable even into your normal daily living and to and to incorporate the activity in into into that".<br><br>PwP (L1 Incomplete, Male): "Because, I mean, you think most of the time you've been totally motivated to get back to it, but then the length of time that you've left it, it's sort of a distant memory because you've not kept on it. And I think that's some of the thing with good habits is you've really gotta keep working at it and not letting it slip".<br><br>PwP (T4 complete, male): "If you can make doing something part of your routine a habit, a good habit. Then you're more motivated to keep it up".<br><br>PwP (T1 Incomplete, Male): "But I liked your idea about building exercise into the things you can do in your life rather than going down the gym. Cause also you're gonna stand out in the gym. "What? What?" You know, there'll be all these pumping people, do you know, pumping iron, and they just look at you and you might upset them".<br><br>PwP (T1 Incomplete, Male): "Now, this meeting has suddenly made me think "there are things I could do". So, the researchers, you've done it. I'm thinking "Hang on a minute. I could pick some bottles up, pick up a can of beans up, do a bit of work while sitting on the sofa"".<br><br>PwP (T4 Complete, Male): "And yeah, I would say that at first, someone might need a need a high level of intervention. Yeah, but then that intervention might become less and less, which it should do as their mind becomes more down to them". |

| | | | | |
|---|---|---|---|---|
| | | | Boredom, being a chore or lack of enjoyment (barrier) (n = 6) | HCP (Physiotherapist, Female): "I think it's really boring and very few of them do it [their prescribed exercise programme] in spite of very specific recommendations and advice around it. But that's exercising rather than not being sedentary".<br><br>HCP (Physiotherapist, Female): "Yeah, just another idea. I've no idea if that's even possible, but just another potential option, leading on from the things that we've said in terms of mood and psychology and exercise and activity, and individualising it as much as possible. Potentially, if you've got different options for different people, they can choose what works for them".<br><br>PwP (T6 Complete, Male): "I think something. Yeah. Yeah, I get told to do something. Sitting and start off from day one, you do, like, as a regime. Day two is still that. Day 3. And by day 25 you're thinking "I'm doing this, this and this" and it all goes out the window. So a year down the line after being told to do this every day".<br><br>PwP (T4 Complete, Male): "There's a thing, isn't it? So, you know, I've got a pill reminder app, right? So it's just says, yeah, "time to take your meds". And it reminds me every like 5 minutes, like "you haven't taken them" and then you go "Oh yeah", and it's actually turned that notification off, where you go "great I've done it now" and it's off. It's not gonna remind me again. Same with exercise".<br><br>PwP (T12 incomplete, Male): "They want you in it [standing frame] for like 3 or 4 hours a day. What can you do, stood? What can you do? It's boring".<br><br>PwP (L1 Incomplete, Male): "Or time slash boredom. As in duration of how long you're motivated. Because it goes back to what we were saying about doing exercises or doing doing things as you probably should when you've left rehab, and then |

| | | | | |
|---|---|---|---|---|
| | | | | over time you've got a bit more lackadaisical about it and bad habits creep in and slip. And we just think "oh I don't need to do that anymore"".<br><br>PwP (L1 incomplete, male): "It [physical activity] being a chore and that might break your motivation cause because it's. It's not enjoyable".<br><br>PwP (T4 Complete, Male): "But there's the thing that cuts across time and boredom, it's enjoyment isn't it. It has to be fun. If it's fun, then all of those things kind of fall into place".<br><br>PwP (T6 Complete, Male): "Otherwise it just becomes the repetitive and that's when you kick it into touch because you can't be bothered [if there isn't choice or variety in an activity routine]".<br><br>PwP (T4 Complete, Male): "There's a balance isn't there. You've got to have enough choice to mix things up".<br><br>PwP (T4 Complete, Male): "Fun, fun tackles prioritising it, other commitments, boredom, time, becoming a chore. And if something is fun to do, you'll do it more often". |
| | | Goals | Goal setting (facilitator) (n = 8) | HCP (Physiotherapist, Female): "It's timing it right, because I think just working out how to live when you first get home, is enough without a goal, because it's such a big shock to the system. But maybe there's a time point after getting home when you want to have goals again. But I don't. If if you said 'you're going home and here are your goals', they might be like 'hold on. I just need to get into my house and work out how to be in my house and doing my routine at home'. It might not fit early in that".<br><br>HCP (Physiotherapist, Female): "The the readmissions, the readmissions, often, so, we see people across the course of |

| | | | | |
|---|---|---|---|---|
| | | | | their lives. But first episode of rehab. So the people that are having an inpatient rehab cycle for the first time following their spinal cord injury, I, like, there is no, there is no service level mechanism for setting goals for post-discharge because, and there might be the odd occasions where you and your patient would talk about something that you thought you know. In fact, no, that's not true".<br><br>PwP (L1 Incomplete, Male): "So if all of these users are on this app, they're all spinal cord injury patients. They're all trying to be more active and less sedentary, and I don't know if, it's not like goal setting, or it's like "Mark's smashed this target" or "he's achieved this this week" and "so and so on has achieved"".<br><br>PwP (L1 incomplete, male): "Realistic goals I guess, that's the one I mean. Because you know, if you start out thinking "Why am I not at the top of the Mount Everest, yet?'"."<br><br>PwP (T6 Complete, Male): "It goes to the old thing about having a beginners, intermediate and an advanced thing".<br><br>PwP (L1 Incomplete, Male): "Is it? I mean, would there be like a mentoring or coaching? It could be cut part of the creating targets. Maybe you can check back or um. Adjustable".<br><br>PwP (T1 Incomplete, Male): "The other thing is maybe you could do like they have in the scouts you could have little badges for can you move your hands? Yes, or no? and then you could do little grip exercises and start from the bottom and work way up. Now, people like Tony who have got a bit more go in them can work their way up to the top. If you start at the bottom and say, little, you know, little bit, baby steps. You start your way at the bottom and work your way up. That's a good training exercise". |

| | | | | |
|---|---|---|---|---|
| | | | | PwP (L1 Incomplete, Female): “Hmm. Yeah. And I mean, Fitbit does the opportunity that you can, when you're tracking your steps, you can do challenges like the number of steps to climb Kilimanjaro, for example, or go from one place to another place. So that's another opportunity. The number of calories you've burnt is enough calories to have done this thing”.<br><br>PwP (T11 incomplete, female): “When I say objectives and goals, I mean personally I like to work towards goals and objectives”.<br><br>PwP (T4 Complete, Male): “I think that's very unique to the specific person and what that person needs are me personally, I wouldn't need that. Some people might need that and therefore I can be a friend, a sponsor or something like that, and they can help together to do it until that person can do it on their own, if that makes sense. So have someone to support you to do it until you can do it on your own”. |
| | | Reinforcement | Rewards (facilitator) (n = 5) | PwP (T12 Incomplete, Male): “Money or vouchers would be wicked [as an incentive for breaking up sedentary behaviour with physical activity]”.<br><br>PwP (L1 Incomplete, Male):<br>I was gonna say a tie in with sponsors. You know, I mean sports shops or sports brands or something, because it's exercise equipment. Or just, yeah, discount, or if you, I don't know”.<br><br>PwP (L1 Incomplete, Male): “Self-esteem. The self-esteem thing might come, you know, if you've readjusted the goals and they’re more realistic you, you're gonna feel a boost. That you're trying again, or you're trying in a different way. So, I don’t know if that answers that one or not”. |

| | | | | |
|---|---|---|---|---|
| | | | | PwP (T4 Complete, Male): “Oh yeah, I think. Anyway, for me personally, I certainly have to reduce and watch what I eat. And I mean, I cycle every day on a day for an hour and a half. Yeah. And things like that. Just so I can basically eat cake and go out of coffee shops. Yeah. I mean, I like eating cake and chocolate, but I do that and the trade-off for me in that is I do three hours of exercise every day, day in, day out. So I can get away with eating, coffee and walnut cake. Because if I did do the exercise, I wouldn't be able to do it. Well, I could”.<br><br>PwP (L1 incomplete, female): “If I could earn a badge for doing a 5-minute exercise on a bad day that might incentivise me for doing it”.<br><br>PwP (L1 Incomplete, Female): “Yeah, I usually do so like, if I go down to the hospital because I've been out for the day and I've done a lot during the day, I'll come back by McDonald's and have a Mcflurry. That's my reward”.<br><br>PwP (T4 Complete, Male): “Well, if if I'm going out seeing friends in coffee shops and now I'd like to have a cake. So you can either have a slab of cake and put a fair bit of weight on, or you can say, you know what? I'm gonna have that cake. So therefore I've burnt the calories and now and exercise just becomes a routine”.<br><br>PwP (L1 Incomplete, Female): “Gems kind of things and you when you get so many gems you get a badge. So it's like a reward that you, “oh, I've. I've got so many. So many from doing so many lessons”. So it kind of motivates you to do that number of lessons because then you've got another reward and it's not much, but it's something”.<br><br>PwP (T11 incomplete, female): “The reward itself would be me completing the goal”. |

| | | | | |
|---|---|---|---|---|
| | Automatic motivation | Emotions | Low mood (barrier) (n = 3) | HCP (Physiotherapist, Female): "I think some people think they've got enough on, you know, like we're all aware of the recommendations with regards to levels of activity that we should have in order to be healthy, but that that advice is even harder to make stick for a population with disabilities sometimes, because they feel like they've been served enough challenge".<br><br>HCP (Physiotherapist, female): "The biggest barrier to general levels of physical activity is how you're thinking and feeling and that comes down to psychology".<br><br>PwP (L1 Incomplete, Male): "Having two daughters and a wife, sedentary behavior is just not allowed, which is good, which is good. They they never allow me to wallow myself in self-pity or lay on the sofa".<br><br>PwP (L1 incomplete, female): "I have good days and bad days and on a good day I'm fine. I can do stuff. It's not a problem, but on a bad day it can vary between not getting out of bed or getting out of bed but not going out the house". |

CCG, Community Caregiver; COM-B, Capability, Opportunity and Motivation to change Behaviour; HCP, Healthcare Professional; PwP, Participant with Paraplegia; TDF, Theoretical Domains Framework.
